# The Healthy Life in an Urban Setting (HELIUS) study in Amsterdam, The Netherlands: cohort update 2024 and key findings

**DOI:** 10.1101/2024.07.16.24310494

**Authors:** Henrike Galenkamp, Anitra D.M. Koopman, J. Esi van der Zwan, Bert-Jan H. van den Born, Anja Lok, Eric P. Moll van Charante, Maria Prins, Arnoud P. Verhoeff, Aeilko H. Zwinderman, Karien Stronks

## Abstract

The Healthy Life in an Urban Setting (HELIUS) study is an ongoing prospective multi-ethnic cohort study, in Amsterdam, The Netherlands that started in 2011. The principle aim of the HELIUS study is to investigate the causes of (the unequal burden of) diseases across ethnic groups, with emphasis on mental disorders, cardiovascular disease and infectious disease, and their interrelationships. Stratified sampling by ethnic group was used to allow for an equal representation of the largest ethnic groups resulting in similar-sized samples of individuals of Dutch, African Surinamese, South-Asian Surinamese, Ghanaian, Turkish and Moroccan origin. A total of 24,780 individuals participated in the baseline examination that consisted of a questionnaire, physical examination and collection of biological material. Follow-up data have been collected through linkage with health care registries and a first follow-up data collection that took part between 2019 and 2022 and included 11,035 participants with an average follow-up time of 6.4 years. The data collection included information on demographics, medical history (including medication use and mental health status), anthropometrics, and fasting blood, urine and stool samples. Here we give an update on the HELIUS study and its methods regarding the first follow-up data collection, data linkage, and additional analyses using stored biomaterials. In addition, we provide a summary of key findings.

## Introduction

Ethnic inequalities in health and health care are widespread and persistent. Societal focus on this inequity has increased over the past decade, in policy, prevention, clinical care, education and research (1, 2). Although the majority of studies on health inequalities between ethnic groups have been performed in the USA, several have also been performed in Europe (3, 4). A growing body of research exists that aims to disentangle the causes of health inequalities, but they remain not fully understood. In general, research on ethnic health inequalities is hampered by a lack of data on ethnic minority groups, either due to exclusion of individuals from scientific studies based on ethnic background, or to a lack of data on ethnic background within studies or medical registrations (5).

In 2010, the Healthy Life in an Urban Setting (HELIUS) study was designed to provide a knowledge base on inequalities in health and health care between the six largest ethnic groups living in Amsterdam, the Netherlands, with a focus on cardiovascular diseases, infectious diseases and mental disorders. HELIUS was set up by the Amsterdam UMC (previously the Academic Medical Center, Amsterdam) and the Public Health Service (GGD) of Amsterdam as a prospective population-based study with stratified sampling across ethnic groups to allow for equal sample sizes. During the baseline data collection between 2011 and 2015, the HELIUS study included nearly 25,000 participants aged 18 to 70 years (average 43.8 years), with 81% being of non-Dutch origin.

Strengths of the study include the inclusion of a large number of migrants and their offspring from several ethnic groups living in the same city, and the collection of a wide range of multi-domain measurements. These study characteristics enable the investigation of ethnic inequalities for several health outcomes, and a wide range of explanatory factors that represent the causal pathway through which ethnicity is linked to incidence and prognosis of health conditions (6). Furthermore, by focusing on three disease categories, potential crosslinks between these diseases can be studied.

The aim of this paper is to provide an update on the yield of the data collection, the first follow-up measurement and additional analyses performed by linkage to health care registries. Particularly, we focus on 1) the first follow-up measurement in 2019-2022, 2) on additional analyses using stored biomaterial, and 3) on additional data collection using registry linkages. We give an overview of the key findings and discuss the implications of HELIUS for clinical practice, health prevention and promotion and provide a brief outline for future research.

## Baseline measurement (HELIUS-1; 2011-2015): Design and response

Between January 2011 and December 2015, baseline HELIUS data were collected among Amsterdam residents of Dutch, Surinamese, Ghanaian, Turkish and Moroccan ethnic origin. People in the age range of 18–70 years were randomly sampled, stratified by ethnic origin, through the municipality register of Amsterdam. This register contains data on country of birth of citizens and of their parents, thus allowing for sampling based on the widely accepted Dutch standard indicator for ethnic origin. This country of birth indicator of ethnicity has the advantage of being objective and stable over time, and cross-validation studies showed a high correlation between the country of birth indicator and self-identified ethnic group indicator among Turkish, Moroccan and Surinamese people in the Netherlands (7). A person was defined as of non-Dutch ethnic origin if he/she fulfilled one of two criteria: (1) he/she was born outside the Netherlands and has at least one parent born outside the Netherlands (first generation) or (2) he/she was born in the Netherlands but both parents were born outside the Netherlands (second generation). For the Dutch sample, we invited people who were born in the Netherlands and whose parents were born in the Netherlands. After data collection, participants of Surinamese ethnic origin were further classified according to self-reported ethnic origin (obtained by questionnaire) into ‘African’, ‘South-Asian’, or ‘other’. Further information on the definition of ethnicity and the stratification that was used in the HELIUS study was previously published (6, 8).

Participants provided written informed consent prior to participation. At baseline, participants were asked permission (1) to store biological samples in the HELIUS biobank for future research (93% agreed), (2) to link their individual data to registries containing data relating to the participants’ health (including hospital admissions, pharmacy data, and vaccination programmes; 90% agreed), (3) to request the official causes of death from Statistics Netherlands (87% agreed) and (4) to approach them for additional studies in the future (substudies; 92% agreed). This enabled us to obtain new laboratory measures from stored samples at baseline and to link HELIUS baseline data to follow-up data (risk factors, health outcomes and mortality) derived from existing registrations.

Data were collected by questionnaire (paper or online, depending on participants’ preference) and a physical examination in which biological samples were also obtained. Ghanaian and Turkish participants unable to fill out questionnaires in Dutch were offered questionnaires in English and Turkish, respectively. All participants unable to complete a questionnaire were offered assistance from an ethnically matched, trained interviewer.

The baseline sample consisted of 24,780 persons (57% women), corresponding to a participation rate of 50% (defined as the share that agreed to participate, out of all persons that we were able to contact and get a response from) and a response rate of 28% (defined as the share that we have baseline data from, out of all persons that were invited). Of the 24,780 participants, 23,936 participants completed the questionnaire, 23,006 completed the physical examination including the collection of biological samples (morning urine, fasting blood and feces) and 22,162 participants completed both.

In the initial HELIUS cohort profile (8), characteristics and methods of the baseline sample are depicted. For example, mean baseline age varied between 39.7 years for Moroccans and 47.6 years for African Surinamese; percentage of first generation migrants varied between 66.6% for Moroccans and 94.4% for Ghanaians, while the percentage with low educational level varied between 3.3% for those of Dutch descent to 31.1% for individuals of Turkish descent.

Participants were asked whether they had family members aged 18–70 years old, who could also be invited. In total, for 8,355 participants we have information regarding family relations with other participants in HELIUS (parent, sibling, child, or partner). This multigenerational design enables us to study both family relations as well as different migration generations.

The HELIUS study is being conducted in line with the Declaration of Helsinki, and has been approved by the Ethical Review Board of the Amsterdam UMC, location AMC.

## Baseline measurements (HELIUS-1): Laboratory measurements on stored samples

During baseline examination, ancillary fasting blood and morning urine samples were collected and stored in the HELIUS biobank. In addition, in sub samples we collected feces samples and vaginal swabs, amongst others. Additional measurements and analyses that have been performed on these stored samples are listed in Table 1.

**Table 1.**
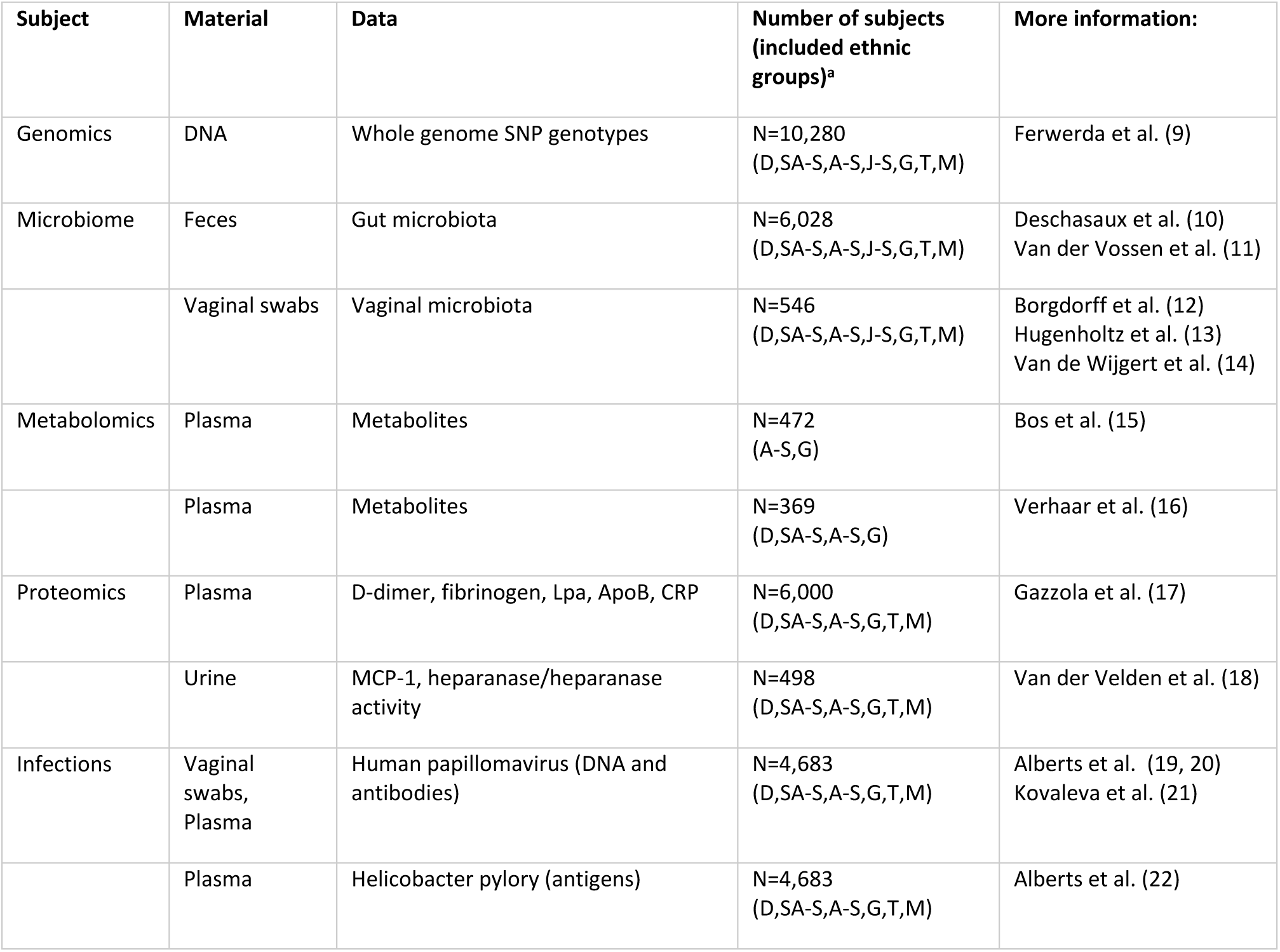
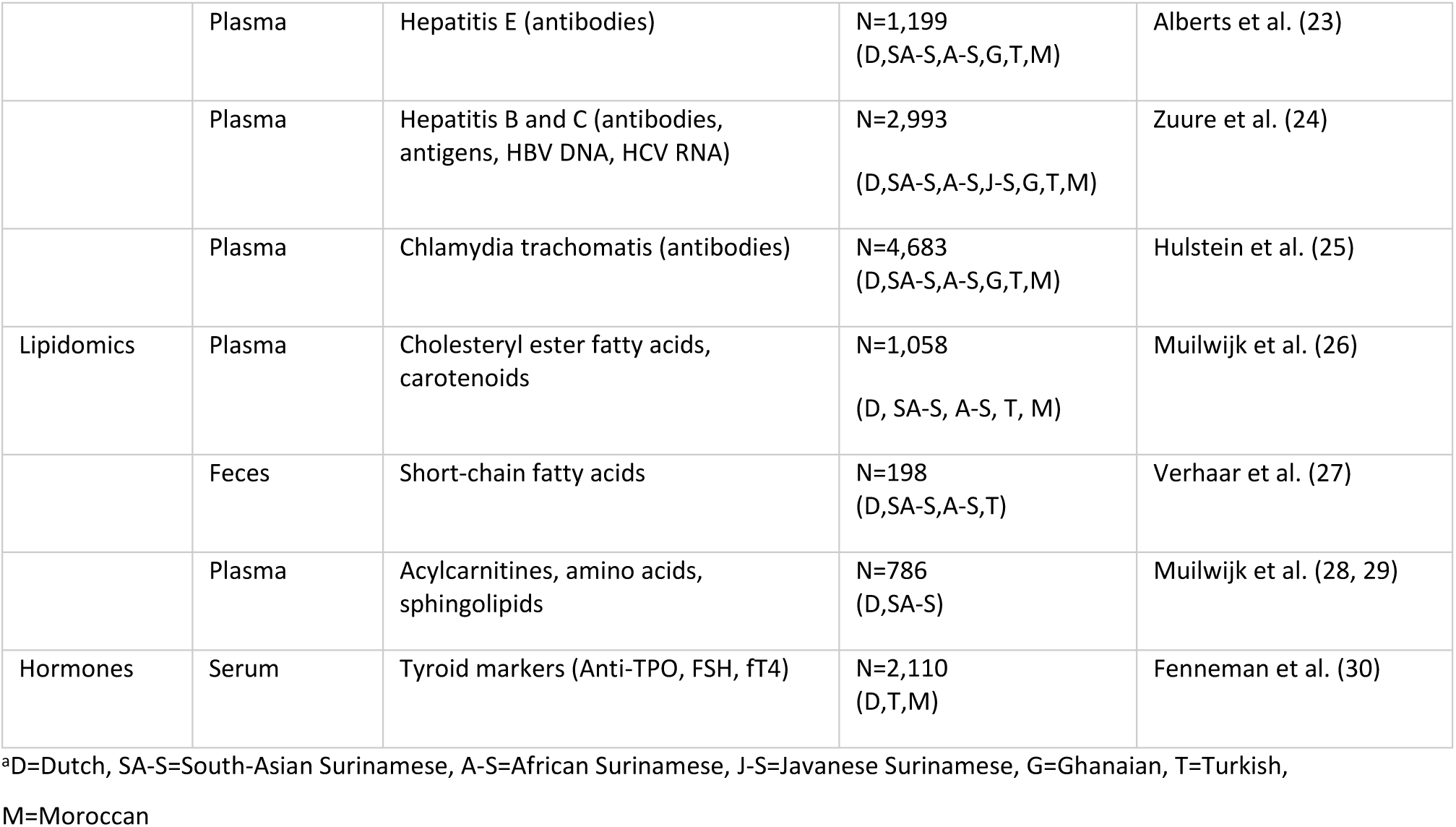
Available data derived from analysis on biological samples collected during HELIUS baseline (2011–2015).

| Subject | Material | Data | Number of subjects (included ethnic groups)<sup>a</sup> | More information: |
| --- | --- | --- | --- | --- |
| Genomics | DNA | Whole genome SNP genotypes | N=10,280<br>(D,SA-S,A-S,J-S,G,T,M) | Ferwerda et al. (9) |
| Microbiome | Feces | Gut microbiota | N=6,028<br>(D,SA-S,A-S,J-S,G,T,M) | Deschasaux et al. (10)<br>Van der Vossen et al. (11) |
|  | Vaginal swabs | Vaginal microbiota | N=546<br>(D,SA-S,A-S,J-S,G,T,M) | Borgdorff et al. (12)<br>Hugenholtz et al. (13)<br>Van de Wijgert et al. (14) |
| Metabolomics | Plasma | Metabolites | N=472<br>(A-S,G) | Bos et al. (15) |
|  | Plasma | Metabolites | N=369<br>(D,SA-S,A-S,G) | Verhaar et al. (16) |
| Proteomics | Plasma | D-dimer, fibrinogen, Lpa, ApoB, CRP | N=6,000<br>(D,SA-S,A-S,G,T,M) | Gazzola et al. (17) |
|  | Urine | MCP-1, heparanase/heparanase activity | N=498<br>(D,SA-S,A-S,G,T,M) | Van der Velden et al. (18) |
| Infections | Vaginal swabs, Plasma | Human papillomavirus (DNA and antibodies) | N=4,683<br>(D,SA-S,A-S,G,T,M) | Alberts et al. (19, 20)<br>Kovaleva et al. (21) |
|  | Plasma | Helicobacter pylori (antigens) | N=4,683<br>(D,SA-S,A-S,G,T,M) | Alberts et al. (22) |
|  | Plasma | Hepatitis E (antibodies) | N=1,199<br>(D,SA-S,A-S,G,T,M) | Alberts et al. (23) |
|  | Plasma | Hepatitis B and C (antibodies, antigens, HBV DNA, HCV RNA) | N=2,993<br>(D,SA-S,A-S,J-S,G,T,M) | Zuure et al. (24) |
|  | Plasma | Chlamydia trachomatis (antibodies) | N=4,683<br>(D,SA-S,A-S,G,T,M) | Hulstein et al. (25) |
| Lipidomics | Plasma | Cholesteryl ester fatty acids, carotenoids | N=1,058<br>(D, SA-S, A-S, T, M) | Muilwijk et al. (26) |
|  | Feces | Short-chain fatty acids | N=198<br>(D,SA-S,A-S,T) | Verhaar et al. (27) |
|  | Plasma | Acylcarnitines, amino acids, sphingolipids | N=786<br>(D,SA-S) | Muilwijk et al. (28, 29) |
| Hormones | Serum | Tyroid markers (Anti-TPO, FSH, fT4) | N=2,110<br>(D,T,M) | Fenneman et al. (30) |

<sup>a</sup>D=Dutch, SA-S=South-Asian Surinamese, A-S=African Surinamese, J-S=Javanese Surinamese, G=Ghanaian, T=Turkish, M=Moroccan

## First follow-up (HELIUS-2; 2019-2022): Design and response

Between May 2019 and November 2022 all baseline participants who were still alive according to the municipality population register and living in The Netherlands were invited for the first follow-up measurement (HELIUS-2). Response rates are presented in Table 2. A total of 371 participants had deceased and 640 participants had moved abroad, leaving 23,769 participants who were eligible for the follow-up measurement. Eligible participants received a written invitation with a response card, and a reminder after four weeks. In case of no response, participants from whom a phone number was collected during the baseline data collection were approached by phone. After five unsuccessful attempts, participants were approached one last time by email (in case an email address was collected during the baseline data collection). After a positive response, participants were contacted by phone by the research team to make an appointment for a visit to the research location. A visit to the research location consisted of a questionnaire/interview, physical examination and collection of biological samples.

**Table 2.**
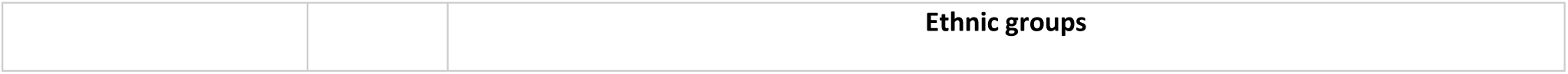
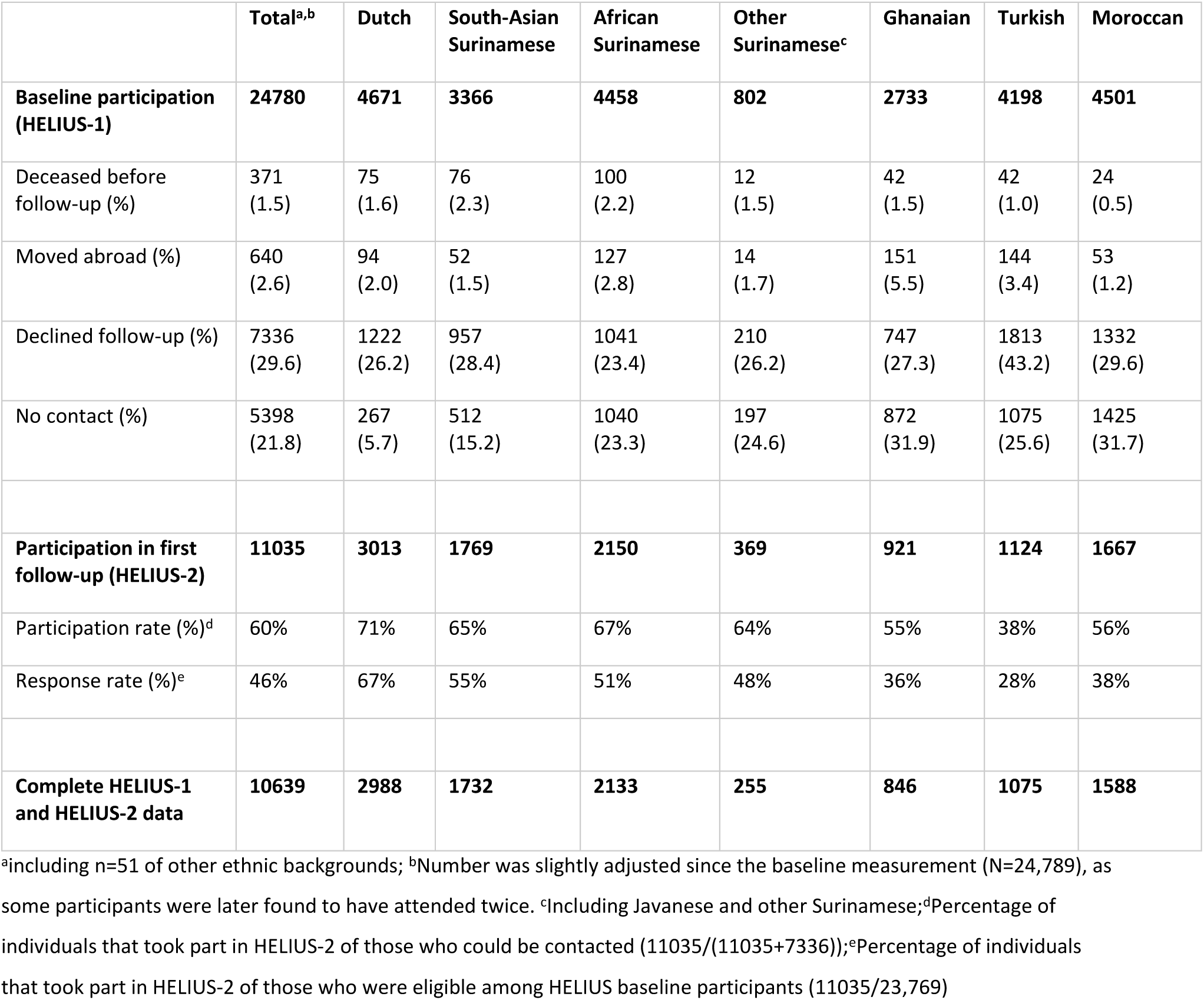
Response rates of the first follow-up of HELIUS (2019-2022; Amsterdam, The Netherlands), by ethnic group.

|  | **Total<sup>a,b</sup>** | **Dutch** | **South-Asian**<br>**Surinamese** | **African**<br>**Surinamese** | **Other**<br>**Surinamese<sup>c</sup>** | **Ghanaian** | **Turkish** | **Moroccan** |
| --- | --- | --- | --- | --- | --- | --- | --- | --- |
| **Baseline participation (HELIUS-1)** | **24780** | **4671** | **3366** | **4458** | **802** | **2733** | **4198** | **4501** |
| Deceased before follow-up (%) | 371<br>(1.5) | 75<br>(1.6) | 76<br>(2.3) | 100<br>(2.2) | 12<br>(1.5) | 42<br>(1.5) | 42<br>(1.0) | 24<br>(0.5) |
| Moved abroad (%) | 640<br>(2.6) | 94<br>(2.0) | 52<br>(1.5) | 127<br>(2.8) | 14<br>(1.7) | 151<br>(5.5) | 144<br>(3.4) | 53<br>(1.2) |
| Declined follow-up (%) | 7336<br>(29.6) | 1222<br>(26.2) | 957<br>(28.4) | 1041<br>(23.4) | 210<br>(26.2) | 747<br>(27.3) | 1813<br>(43.2) | 1332<br>(29.6) |
| No contact (%) | 5398<br>(21.8) | 267<br>(5.7) | 512<br>(15.2) | 1040<br>(23.3) | 197<br>(24.6) | 872<br>(31.9) | 1075<br>(25.6) | 1425<br>(31.7) |
| **Participation in first follow-up (HELIUS-2)** | **11035** | **3013** | **1769** | **2150** | **369** | **921** | **1124** | **1667** |
| Participation rate (%)<sup>d</sup> | 60% | 71% | 65% | 67% | 64% | 55% | 38% | 56% |
| Response rate (%)<sup>e</sup> | 46% | 67% | 55% | 51% | 48% | 36% | 28% | 38% |
| **Complete HELIUS-1 and HELIUS-2 data** | **10639** | **2988** | **1732** | **2133** | **255** | **846** | **1075** | **1588** |

<sup>a</sup>including n=51 of other ethnic backgrounds; <sup>b</sup>Number was slightly adjusted since the baseline measurement (N=24,789), as some participants were later found to have attended twice. <sup>c</sup>Including Javanese and other Surinamese; <sup>d</sup>Percentage of individuals that took part in HELIUS-2 of those who could be contacted (11035/(11035+7336)); <sup>e</sup>Percentage of individuals that took part in HELIUS-2 of those who were eligible among HELIUS baseline participants (11035/23,769)

Of all eligible baseline participants, 7,336 declined participation in the first follow-up, and 5,398 could not be reached. In total, 11,035 persons took part in HELIUS-2, of whom 10,639 had a complete baseline examination (completed questionnaire and physical examination). Response rates (HELIUS-2 participants/eligible participants) varied from 28% in the Turkish origin group to 67% in the Dutch origin group. When taking into account that not all participants could be reached, participation rates (HELIUS-2 participants/eligible participants who were reached) varied from 38% in the Turkish origin group to 71% in the Dutch origin group.

Distribution of demographic characteristics of the follow-up sample is shown in Table 3. In this table, only the six largest ethnic groups are shown separately, because of a low number of participants in the other Surinamese (n=369) and other origin (n=22) groups. In all six ethnic groups, women took part in the first follow-up more often than men, and participants were most often aged between 50-59. Most participants were migrants themselves (first generation, range 74-96%), as compared to offspring of migrants (second generation, range 4-27%). Educational level was only measured at baseline, and varied across ethnic groups. On average, and this was reflective of the actual situation of the Amsterdam population, Dutch origin participants had the highest educational level, and Ghanaian origin participants the lowest (8).

**Table 3.**
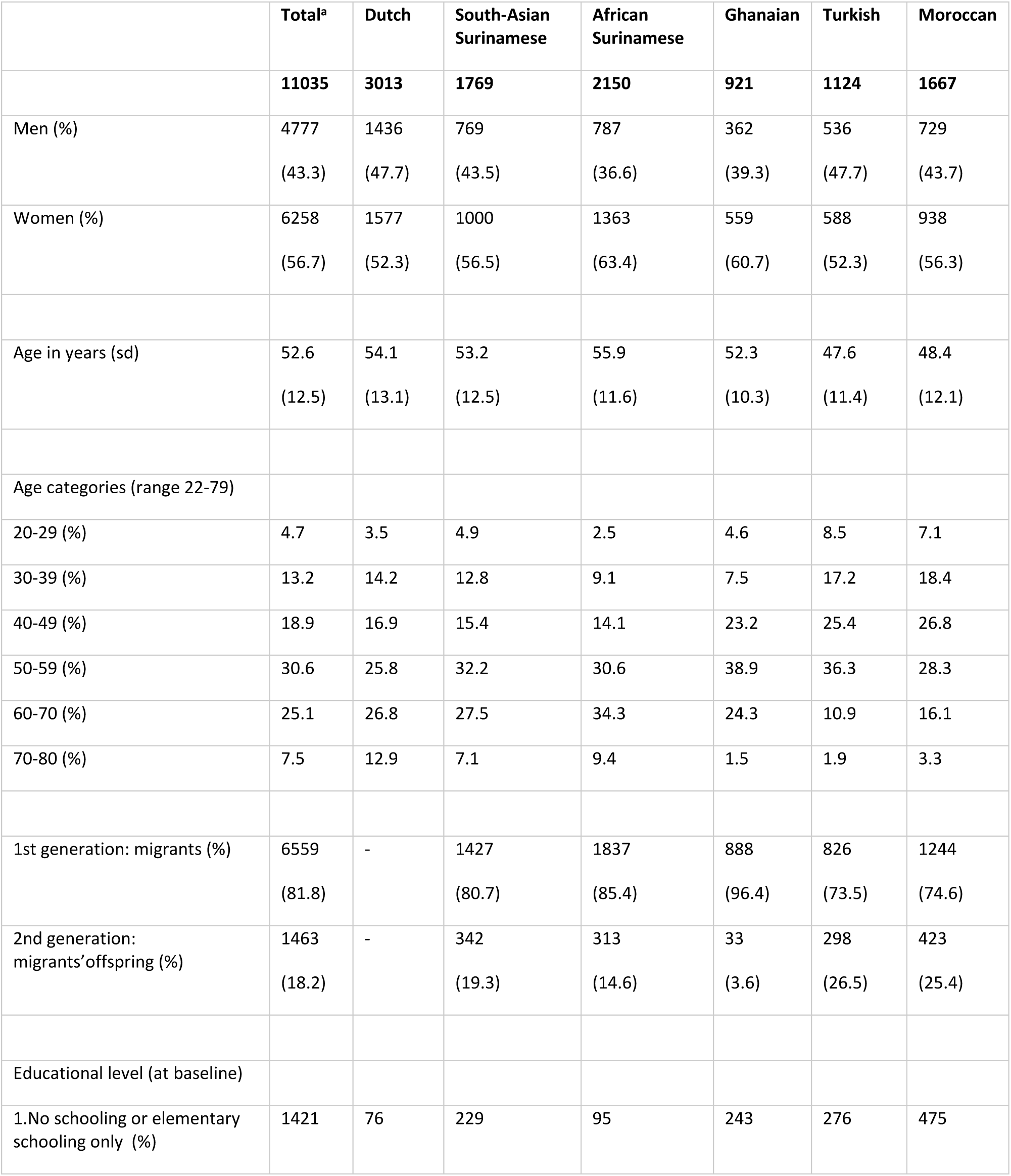
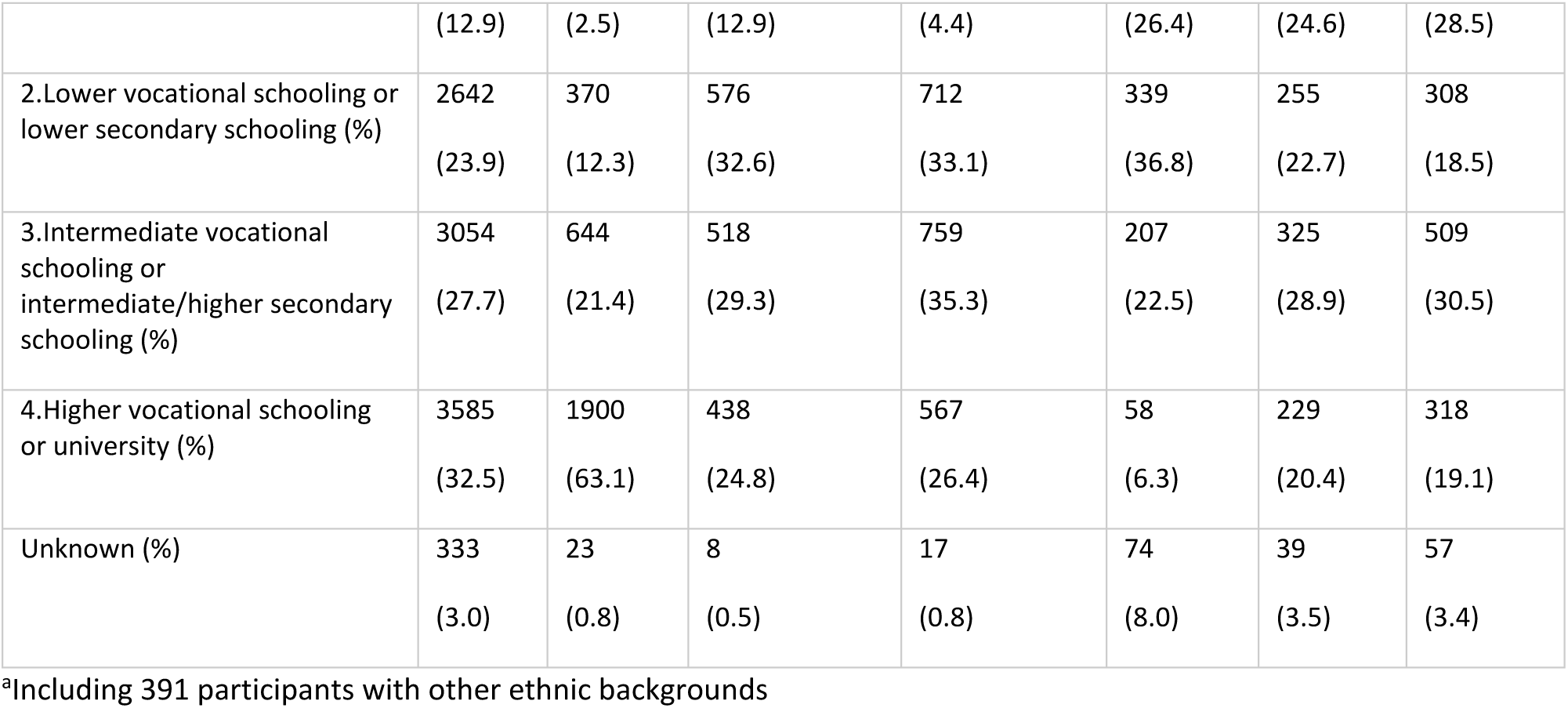
Demographic characteristics of HELIUS-2 participants, by ethnic group.

Within the total HELIUS baseline population, it appeared that those who participated in the follow-up data collection were generally older than those who did not, more often first generation migrants, and more often had intermediate or higher educational levels (Supplemental Table 1, Supplemental Figure 1).

Regarding health status (Table 4), in most ethnic groups participants of the first follow-up less often had diabetes or self-reported cardiovascular disease, compared to those HELIUS participants who did not participate in the first follow-up. In addition, they smoked less often and met the physical activity guideline more often. Finally, participants included at follow-up reported less often depressive symptoms at baseline. After excluding participants who deceased or moved abroad between HELIUS-1 and HELIUS-2, these differences between participants and non-participants remained but were attenuated (Supplemental Table 2).

**Table 4.** Comparison of selected baseline health characteristics between participants who completed follow-up and those who did not.

|  |  | Total<sup>a</sup> | Dutch | South-Asian<br>Surinamese | African<br>Surinamese | Ghanaian | Turkish | Moroccan |
| --- | --- | --- | --- | --- | --- | --- | --- | --- |
| **Diabetes (%)** | H2 | 9.3 | 3.2 | 17.9 | 10.1 | 9.8 | 8.0 | 10.8 |
|  | No H2 | 12.1 | 4.4 | 21.6 | 14.0 | 12.1 | 11.0 | 11.9 |
|  | p-value<sup>b</sup> | <.001 | 0.047 | 0.012 | <.001 | 0.078 | 0.006 | 0.314 |
| **Hypertension (%)** | H2 | 37.3 | 29.6 | 41.1 | 50.7 | 54.4 | 27.5 | 26.2 |
|  | No H2 | 37.4 | 29.9 | 44.4 | 50.4 | 55.4 | 30.0 | 23.0 |
|  | p-value | 0.848 | 0.865 | 0.075 | 0.852 | 0.648 | 0.134 | 0.021 |
| **Self-reported CVD (%)** | H2 | 4.5 | 3.1 | 7.5 | 4.9 | 4.0 | 5.5 | 3.3 |
|  | No H2 | 6.3 | 5.2 | 11.9 | 6.1 | 5.0 | 7.3 | 3.4 |
|  | p-value | <.001 | <.001 | <.001 | 0.087 | 0.264 | 0.040 | 0.930 |
| **Current smoker (%)** | H2 | 21.1 | 22.0 | 24.0 | 27.9 | 4.9 | 29.6 | 10.3 |
|  | No H2 | 28.1 | 30.8 | 37.9 | 37.8 | 4.0 | 37.8 | 16.1 |
|  | p-value | <.001 | <.001 | <.001 | <.001 | 0.297 | <.001 | <.001 |
| **Alcohol use (intermediate/high) (%)<sup>c</sup>** | H2 | 27.1 | 64.8 | 15.7 | 18.3 | 11.5 | 8.2 | 3.3 |
|  | No H2 | 17.1 | 60.7 | 18.1 | 20.5 | 10.0 | 6.0 | 3.2 |
|  | p-value | <.001 | 0.006 | 0.064 | 0.068 | 0.269 | 0.015 | 0.860 |
| **Meeting Dutch physical activity guideline (%)** | H2 | 59.6 | 75.5 | 53.1 | 61.3 | 54.7 | 43.3 | 47.8 |
|  | No H2 | 46.3 | 72.1 | 42.9 | 52.7 | 47.7 | 34.9 | 38.7 |
|  | p-value | <.001 | 0.012 | <.001 | <.001 | 0.001 | <.001 | <.001 |
| **Depressive symptoms (PHQ-9 sum score ≥10) (%)** | H2 | 12.1 | 6.0 | 17.3 | 9.2 | 8.8 | 20.5 | 18.7 |
|  | No H2 | 17.0 | 9.6 | 20.0 | 11.8 | 8.9 | 23.9 | 21.6 |
|  | p-value | <.001 | <.001 | 0.045 | 0.005 | 1.00 | 0.023 | 0.023 |

<sup>a</sup>including other ethnic backgrounds; <sup>b</sup>chi-square test (2 sided exact test); <sup>c</sup>Intermediate/high (men >4 glasses/week, women >2 glasses/week)
H2: participation in HELIUS-2; No H2: no participation in HELIUS-2 (either because of mortality, relocation to other country, refusal, or no contact)

## First follow-up: Measurements in total sample

The measurements in the first follow-up followed the same methods as those of the baseline data collection. While HELIUS baseline included a broad range of determinants of health, this follow-up data collection focused on a smaller set of outcome measures, predominantly related to mental health and cardiovascular disease. In addition to that, more in-depth measurements were performed in smaller samples (see next paragraph). An extensive list of measures of the baseline examination can be found in the initial HELIUS cohort profile (8). Table 5 describes the measures that were taken at both measurement waves, with aligned methods between the two rounds as much as possible. An important difference is that at baseline participants filled in the questionnaire (paper or digital version) at home, if needed with help from an interviewer. At follow-up, the questions were asked by the research nurse at the research location. Questionnaires consisted of repeated demographic questions on marital status and work status, but also questions concerning health behaviours, medical history, depressive symptoms, mastery and negative life events. Physical examination included anthropometry and blood pressure measurements. Finally, fasting blood samples were drawn and morning urine and fecal samples were collected for testing. Except for the fecal samples, no HELIUS-2 biological samples were stored in the biobank.

**Table 5.** Overview of measurements that were done in both HELIUS-1 and HELIUS-2, or only in HELIUS-2.

|  | **Baseline**<br>**HELIUS-1**<br>**(2011-2015)**<br>**N~25000** | **Follow-up**<br>**HELIUS-2**<br>**(2019-2022)**<br>**N~11000** |
| --- | --- | --- |
| **Variables questionnaire/interview** |  |  |
| Marital status | X | X |
| Occupational status | X | X |
| Smoking | X | X |
| Alcohol intake | X | X |
| Self-reported history of high blood pressure | X | X |
| Self-reported history of diabetes | X | X |
| Self-reported myocardial infarction | X | X |
| Self-reported dotter/by-pass surgery | X | X |
| Self-reported cerebrovascular events | X | X |
| Mastery (Pearlin-Schooler Mastery Scale) | X | X |
| Depressive symptoms (PHQ-9) | X | X |
| Stressful life events | X | X |
| **Physical examination** |  |  |
| Current medication use (name, frequency, indication) | X | X |
| Anthropometry (weight, height, and circumferences of waist, hip) | X | X |
| Blood pressure (sitting position, 5 min of rest, WatchBP Home, Microlife) | X | X |
| **Laboratory measurements fasting blood** |  |  |
| Glucose, HbA1c, haemoglobin, triglycerides, total cholesterol, HDL, LDL (calculated), creatinine | X | X |
| ASAT, ALAT, GAMMA-GT, Trombocytes |  | X |
| **Laboratory measurements morning urine** |  |  |
| Microalbumin, creatinine | X | X |
| **Faeces (sub samples)** |  |  |
| Fecal microbiome (16S) | X<br>N=6,029 | X<br>N~8000 |
| Stored faeces samples | X<br>N=6,046 | X<br>N~8000 |
| **Swabs (sub samples)** |  |  |
| Nasal and throat swabs | X<br>N=6,631 | X<br>N~2,000 |
| Oral swabs |  | X<br>N~2,000 |

## First follow-up: In-depth measurements in subgroups

In addition to measurements in the total study population of HELIUS-1 and HELIUS-2, specific measurements have been performed in smaller sub groups in between 2020 and 2022. Firstly, between 2020 and 2022 a longitudinal serological study was performed, to study ethnic inequalities in the epidemiology of COVID-19 (31). During three study visits among a subsample of ∼2,500 participants (six largest ethnic groups, random samples), SARS-CoV-2-specific antibodies were determined and an interview was conducted containing questions on COVID-19-related symptoms, reach and uptake of control measures, well-being, risk of exposure (work environment, contacts, traveling) (Supplemental Table 3) and severity of disease. In addition, an online questionnaire was sent to all HELIUS baseline participants for whom we had an email address, and was filled in by 4,450 participants. Questions focused on perceived changes in finances, health behaviors, mental health factors, and use of non-COVID-19 health care due to the COVID-19 pandemic (32). Secondly, in an additional visit after the HELIUS follow-up visit, a Fibroscan was performed in ∼400 participants (all ethnic groups) to measure liver stiffness and liver steatosis (33). Participants were selected based on a high-risk of metabolic dysfunction-associated steatotic liver disease (MASLD), or were included in the control group. Finally, vascular ageing was studied in a subsample of ∼3,000 participants of Dutch, South-Asian Surinamese and Moroccan origin (aged 35-65) through carotid ultrasound examination (34), followed by a carotid and brain MRI (n∼570) and cardiac MRI (n∼150).

## Linkage with health registries and other databases

Next to data collected from participants themselves, data on exposures and outcomes are repeatedly being obtained via linkage with registries. Up until 2022, linkages have been established on an individual level for different health outcomes (including diabetes, CVD (hospitalization), infectious diseases (hospitalization), (all cause-) mortality and cancer). In addition, data on environmental exposures (including safety, food environment, air pollution) were obtained on neighborhood or postal code level. Table 6 provides an overview of the available data.

**Table 6.** Overview of established linkages on health outcomes and exposures.

|  | Type of data | Period | Sample size | Data source | More information: |
| --- | --- | --- | --- | --- | --- |
| Outcomes | Health care use (including ATC drug codes, health care use abroad) | 2010-2015 | N=15,461 | Achmea Health database | Şekercan et al. (35)<br>Schuts et al. (36) |
|  | Hospital admissions | -2020<br>Regularly updated | N=19,893 | Statistics Netherlands, Dutch Hospital Data | Bolijn et al. (37) |
|  | Incident diabetes | 2011-2017 | N=17,829 | Vektis | Muilwijk et al. (29) |
|  | Cancer diagnoses and care | 1989-2019 | N=19,931 | Dutch Cancer Registry | Van Tuijl et al. (38, 39)<br>Müller et al. (40) |
|  | COVID-19 testing and vaccination | March 2020-August 2021 | N=14,589 | Municipal Health Service of Amsterdam | Chilunga et al. (41)<br>Campman et al. (42) |
|  | Causes of death | 2011-2020<br>Regularly updated | N=293 | Statistics Netherlands | Bolijn et al. (37) |
|  | General Practitioner data | -2022 | N=9,212 | Academic GP network AN(H)A | Currently ongoing |
| Exposures | Environment (pollution, SES, noise, livability, etc) | 2011-2015<br>(inclusion date)<br>Regularly updated | N=24,780 | Geoscience and Health Cohort Consortium (GECCO) | Generaal et al. (43)<br>www.gecco.nl |
|  | Food environment | 2011-2015<br>(inclusion date) | N=24,780 | Locatus | Poelman et al. (44) |
|  | Safety data | 2011-2015<br>(inclusion date) | N=20,463 | Safety data O&S | Timmermans et al. (45) |
|  | Pregnancy data | 2000-HELIUS<br>baseline | N=3,034 | Perined | Burger et al. (46) |

## Key findings and publications

A list of all publications that are based on data from the HELIUS study is available at https://www.heliusstudy.nl/nl/researchers/publications. In this section we highlight the key findings up to 2023, based on the conceptual framework that underpins the HELIUS study (as illustrated in Figure 1). This framework constitutes the initial reference point for conducting analyses to elucidate the factors contributing to ethnic disparities in health.

**Figure 1.**
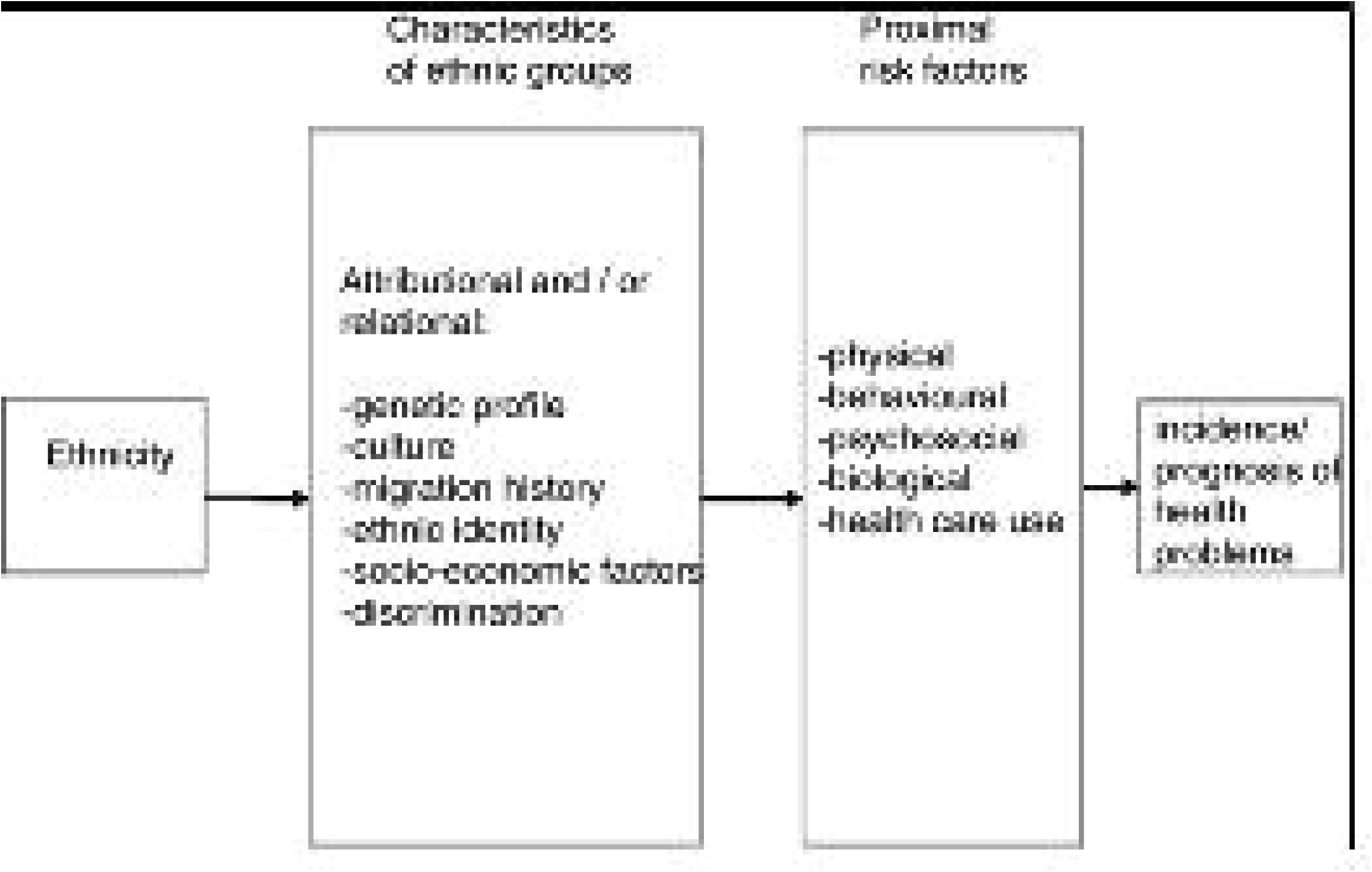
Original conceptual framework integrating possible explanations for the relationship between ethnicity and health. (6)

The model specifies that ethnicity is associated with an uneven distribution of specific risk factors. They are also called proximal factors as they are considered to be more proximate to the onset of pathogenic processes. These include biological (e.g. risk of hypertension), physical (e.g. working conditions), behavioural (e.g. smoking), psychosocial (e.g. stress), and health care use. The uneven distribution of these proximal risk factors across ethnic groups is no coincidence: this distribution is grounded in a context of several specific circumstances. The literature indicates at least the following causal pathways that link ethnicity to health via the proximal risk factors, depicted in the Figure as attributional and/or relational factors: genetic profile, culture, migration history, ethnic identity, socio-economic position, and discrimination. By incorporating the distribution of proximal factors within these causal pathways, scientific research can generate findings explaining ethnic health disparities that are applicable to other ethnic groups sharing similar characteristics. Simultaneously, these causal pathways might also help to explain why health status varies within a certain group.

Findings of the HELIUS study will be categorized by theme: cardiovascular diseases, mental disorders and infectious diseases. In addition, conform the conceptualization in Figure 1, several types of studies can be distinguished: 1) studies that assess ethnic inequalities in risk factors or in health outcomes (either prevalence, incidence, or prognosis of health problems), 2) studies focusing on the association between risk factors and health outcomes, either in the total HELIUS population or stratified by ethnicity (moderation), and 3) studies elucidating how risk factors explain inequalities in health outcomes. All these studies meet the central aim of HELIUS. Studies also often take into account patterns across groups based on demographics other than ethnicity, such as age, sex and education. In addition, HELIUS continuously offers possibilities to 4) examine measurement differences, or differences in reference values for risk factors or outcomes across ethnic groups. Finally, HELIUS studies 5) contribute to meta-analyses, that generally include populations with no or limited diversity.

## Cardiovascular diseases

HELIUS has shown that there are substantial inequalities in cardiovascular health between ethnic groups, particularly in the prevalence of diabetes (47), hypertension (48) and chronic kidney disease (49). One of the most noticeable inequalities includes the prevalence of diabetes, which in general is two to four times more common among individuals with a migration background compared to the Dutch host population, and also has an onset at a much younger age (47). In addition, diabetes-related complications, including coronary artery disease and nephropathy, are more prevalent among migrants, especially among individuals of South-Asian Surinamese, Turkish and Moroccan descent (50). Potential explanatory mechanisms include ethnic differences in the prevalence of hypertension and obesity. However, also in normal weight individuals cardiometabolic risk remains increased (51). HELIUS has shown that the relation between diabetes and measures of body composition, including BMI and waist circumference, differs across ethnic groups (52). This finding from HELIUS and other studies has contributed to ethnicity-specific recommendations for case-finding of type 2 diabetes in the Netherlands (53): for South-Asian Surinamese, case-finding is recommended for those who are 35 years or older, and Turkish and Moroccans when they are 45 years or older, compared to 50 years and older for all other groups. Whether the higher prevalence of obesity and diabetes in individuals with a migration background may relate to ethnicity-specific dietary influences, specific metabolites and differences in gut microbiota composition is currently being studied.

Besides inequalities in the risk of diabetes, HELIUS has also contributed to our understanding of ethnic inequalities in the prevalence of hypertension. Individuals of Ghanaian, African Surinamese and South-Asian Surinamese descent exhibit a higher risk of hypertension compared to other ethnic groups (54), a risk that cannot be fully explained by variations in proximal risk factors such as BMI, physical activity, and attributional/relational factors like educational level (55). However, educational level remains a key determinant of poor blood pressure control, regardless of ethnic background (56). In line with this finding, the recent European guideline on cardiovascular disease prevention recommends multiplying factors to take into account CVD risk imposed by ethnicity, independent of other risk factors. Additionally, recent risk estimations based on national mortality data suggest an increased risk of death from CVD among various ethnic minority groups, potentially warranting lower risk thresholds for cardiovascular screening (57). Moreover, measures aimed at improving health literacy and medication adherence may play a crucial role in helping patients achieve their target values (56, 58). Interestingly, treatment rates were similar or even higher among Ghanaians and African and South-Asian Surinamese compared to the Dutch population (59).

Finally, a recent analysis of data from the baseline and first follow-up measurement has shown that ethnic disparities in the prevalence of hypertension appear to increase over time, especially in those aged ≤ 45 years (60). An increase in overweight among the younger age groups appears to be partly responsible for these growing disparities. Obesity has been shown to substantially contribute to the high cardiovascular risk observed across all ethnic groups (61). Even after accounting for obesity, ethnic disparities in cardiovascular risk and in changes in cardiovascular risk over time persist (51, 60). This underscores the imperative for additional research into the causes of ethnic variations in cardiovascular disease.

## Mental disorders

The majority of studies in the mental health domain have focused on depressive symptoms, which have a higher prevalence in all migrant groups in HELIUS compared to those with Dutch origin, especially in Turkish and South-Asian Surinamese individuals (62, 63). A comparison between migration generations showed that also second generation migrants had an increased risk of depressive symptoms, compared to their Dutch counterparts (63). In addition, ethnic inequalities were observed in substance (ab)use (64–66), post-traumatic stress disorder (PTSD) (67) and suicidal ideation (68). In general these disparities reflect an increased risk in those with a migration background, with the exception of regular alcohol use, which is more common in those with Dutch origin (69). Disparities were also observed in proximal risk factors for depressed mood, including childhood maltreatment (70, 71). The prevalence of any type of childhood maltreatment ranged from 24% in the Moroccan origin group to 39% in the African Surinamese group, with generally higher rates in women than in men. Emotional neglect, the most prevalent type of maltreatment, and psychological abuse were significantly associated with a higher prevalence of depressed mood in all ethnic groups (70). Other proximal risk factors for depressed mood include perceived ethnic discrimination (68, 72, 73), specific dietary patterns, such as patterns that are high in sugar and saturated fat (74–76), and short sleep (77). In addition, socioeconomic factors (63, 78), including unfavourable working conditions (79), and environmental factors (43, 80) were shown to be correlated with depressed mood. It has also been assessed whether coping strategies for mental health might be different across ethnic groups. This appeared not to be the case for mastery, which had a mediating role in the relationship between ethnic discrimination and depressive symptoms, and this was similar across ethnic groups (72).

Regarding the interplay between mental health and cardiometabolic diseases, we found that depressive symptoms and trauma/PTSD were associated with obesity (81), metabolic syndrome (67), and hypertension (82).

Recent studies on the gut-brain-axis were performed. It was shown that diversity of the gut microbiota, as well as specific microbial taxa, are related to depressive symptoms, even after adjustment for socioeconomic factors, lifestyle and medication use. The substantial ethnic differences in depression do indeed appear to be related to ethnic differences in the microbiome (83, 84).

## Infectious diseases

Research on infectious diseases in HELIUS has focused on the epidemiology of human papillomavirus (19), viral hepatitis B, C and E (23, 24), chlamydia (25), helicobacter pylori (22), and, more recently, COVID-19 (31). We found in a subset of participants that Ghanaian (age– and sex-adjusted prevalence of 5.4%), Turkish (4.1%) and African-Surinamese (1.9%) first generation migrants were at increased risk of chronic hepatitis B (HBV) infection and many were unaware of their infection, whereas hepatitis C (HCV) prevalence was low among all ethnic groups (24). These results were used by the Dutch Health Council in their advice to the Minister of Health, Welfare and Sport in the Netherlands to limit screening for these infections to key populations (85, 86), and this advice was adopted (86). Among sexually active participants aged 18-31 years, chlamydia trachomatis prevalence, tested in stored blood samples, was higher among African Surinamese (72%) and Ghanaian (68%) ethnic groups, compared to the group of Dutch origin (38%) (25).

Studies that were set up during the COVID-19 pandemic showed that SARS-CoV-2 incidence was higher in the ethnic minority groups compared to the Dutch origin group. Inequalities became wider during the second wave versus the first wave of infections, which may imply that very stringent measures during the first wave of infections likely prevented ethnic disparities in the initial spread of the virus, but subsequent less rigorous measures might have resulted in a more rapid spread in the ethnic minority groups (31, 87). In line with these findings, we found that the risk of COVID-19 hospitalization was higher in all ethnic minority groups, but the risk of adverse outcomes after COVID-19 hospitalization was similar (88). Additionally, individuals with a migration background and those in lower education or occupation categories were more susceptible to adverse changes across various life domains, including health behavior, mental well-being, and access to non-COVID-19 health care (32).

In general, disparities in infectious disease risk might be explained by a different infection prevalence in countries of origin, different risk of exposure to pathogens in the Netherlands, increased susceptibility, increased rates of co-morbidities and biological differences between ethnic groups. For chlamydia, it appeared that indicators of socioeconomic status, and sexual risk and health seeking behavior could not fully explain ethnic disparities that were found (25). For COVID-19, our results suggested that inequalities in hospitalization risk were also attributable to the higher rate of comorbidities (i.e., diabetes and hypertension) in specific ethnic minority groups. We also found that levels of knowledge regarding antibiotics use and prescriptions varied between ethnic groups (36). This is of concern, as antimicrobial resistance can impact the treatment and prevention of bacterial infections. Finally, we showed a lower intent to vaccinate against SARS-CoV-2 in ethnic minority groups (89). Ethnic-specific determinants (e.g. feeling less emotionally distressed due to COVID-19 in the Dutch group) and general determinants (e.g. being female) of lower vaccination intent observed in our study, could help shape vaccination interventions and campaigns and reduce severe outcomes of SARS-CoV-2 infection.

## Migrants vs migrants’ offspring

Beneath the surface of ethnic health disparities lies the hypothesis that these inequalities may diminish with time, as migrants establish longer-term residence in the Netherlands. Moreover, it is often assumed that the health of migrants’ descendants aligns more closely with the Dutch population in terms of economic status, lifestyle choices, and healthcare utilization. On the other hand, the healthy immigrant paradox reflects that those that migrate are a healthier selection of the total population in the country of origin, showing better health than natives when they arrive in the host country (90). These advantages however disappear over time in the host country. More empirical data is needed to shed light on these hypotheses regarding migrants in the Netherlands.

Studies in HELIUS showed that ethnic inequalities were similar in subgroups of residence duration and acculturation (91), and, in most cases, the generation of migrants’ offspring still exhibits a higher risk of disease and unfavourable risk factor profile, compared to their Dutch counterparts. For example, Helicobacter pylori, one of the most common bacterial infections worldwide, was more often present in blood of first-generation migrants and prevalence remained elevated among their offspring when compared to the Dutch origin group (22). The disease risk might even be higher in migrants’ offspring than in those who migrated themselves: belonging to the group of migrants’ offspring compared to first-generation migrants was significantly associated with more nasal allergy, eczema and asthma (92). In certain instances, aligning with the Dutch population may not represent a desirable shift for individuals of migrant heritage. For example, the gut microbiota of younger generations (both those with and without a migration background) is transitioning towards a less complex and less proficient fermentative configuration, which is associated with diseases of affluence (11). Finally, migrants’ offspring still had an increased risk of depressed mood, compared to the host population (63). This could partly be explained by their worse social conditions (SES, perceived discrimination and sociocultural integration). Thus, HELIUS results so far do not lend support to the hypothesis of decreasing ethnic inequalities across generations.

## Explaining inequalities

In summary, there are significant ethnic differences in the risk of cardiovascular, mental and infectious diseases, which for some diseases seem to persist among migrants’ offspring. While our findings suggest that ethnic minority groups generally face increased risks, no specific group consistently exhibits a greater risk across the broad spectrum of diseases. This emphasizes the need to differentiate between individual ethnic groups.

We found evidence that all categories of proximal factors identified in the initial conceptual model explain some of the disparities in disease risk. Whereas the focus was on behavioural and biological risk factors in particular (e.g. obesity, physical activity, lipid levels), also environmental conditions (e.g. working conditions) and psychosocial factors have been shown to explain part of the worse health of ethnic minority groups. In addition, we found evidence for a differential association between proximal risk factors and health outcomes. We also studied proximal risk factors that have, so far, been less studied among ethnic minority groups, in particular the microbiome. These studies show that diversity of vaginal and gut microbiota composition differs between ethnic groups, and also that the association between microbiome phenotypes and disease outcome varies across ethnic groups (10, 12, 93).

Regarding the characteristics of ethnic groups that link these proximal risk factors to ethnicity, the analyses from the HELIUS study confirmed conclusions from previous research that ethnic inequalities in health are at least partly socio-economic in nature. However, the novelty of the HELIUS analyses lies in the insights generated on other categories of explanatory factors than SES, for example based on the finding that perceived discrimination explained a substantial part of the variation in depressive symptoms (62). Also, group-level determinants such as the food environment and social norms regarding body weight have been shown to be important (94). Finally, all other categories of relational/attributional factors in Figure 1 have been shown to be important, either in explaining inequalities in health *between* the ethnic minority groups and the population of Dutch descent, or in identifying variation in health *within* the ethnic minority populations. Examples of the former include ethnic differences in genetic profile and associations with pro-diabetogenic lipid profile (95) and the gut microbiome (96), and acculturation variables explaining variation in dietary patterns (97) and in body size ideals (98).

Several studies have been performed on the validity and reliability of instruments to measure mental health outcomes, in particular focusing on cross-cultural validity. Examples include validity studies of the SF-12 to measure quality of life (99), the PHQ-9 to measure depressive symptoms (100), the Fagerström test (64), and the Cannabis Use Disorders Identification Test (CUDIT) (65). Results varied per measurement instrument. For example, the PHQ-9 showed to be measurement invariant across ethnic groups (100), whereas differential item functioning needs to be taken into account when using the Fagerström test to assess nicotine dependence across ethnic groups (64). These results can inform other studies and clinical practice regarding the feasibility of using existing instruments in heterogeneous study samples with regard to ethnicity.

An important aim of HELIUS is to contribute to evidence on health outcomes and risk factors in an increasingly diverse population. This objective is primarily pursued through the comparative analysis of the specific ethnic groups encompassed within the HELIUS study. But HELIUS is also increasingly being included in multi-cohort consortia, with the aim to increase the generalizability of results to real-world populations, as opposed to homogeneous populations. Examples include the Biobanking and Biomolecular Resources Research Infrastructure (BBMRI) (15) and the NCD RisC factor collaboration, which brings together data on major risk factors for non-communicable diseases for all countries of the world. In addition, HELIUS participates in several individual-level meta-analyses, such as MoodFood that studies the association between diet and depression (76), a meta-analysis on pathobionts in the vaginal microbiota (14), PSY-CA on the relationship between psychosocial determinants and cancer outcomes (39), and the earlyCause consortium on early-life stress and cardiometabolic outcomes (101).

## Main strengths and weaknesses

Cohort studies require major, recurring research investments. Regarding HELIUS, this has led to a cohort that includes a large sample size of people from different ethnic groups, including those of Dutch origin. In combination with a wide range of health determinants, the study enables a thorough study of inequalities in health in a multi-ethnic population. This is achieved not only through recurring measurement rounds with intervals of several years, but also by leveraging the existing cohort structure to establish sub-studies in a relatively short timeframe. We effectively used this opportunity, for example, during the COVID-19 pandemic. The representativeness of the sample at baseline is another strength, which provides a basis for extrapolating results to the total population (8). At the same time, the relatively limited sample size at follow-up poses a threat to this representativeness, as groups who did and did not participate in the follow-up measurement round differed with regard to demographic and health characteristics. Another limitation is the unknown validity of some of our questionnaires and physical measurements, which may still mask an unknown proportion of health disparities across ethnic groups. Although some measures have been checked for cross-cultural validity, this is not possible with all measures, as gold standards are often not present. Finally, an inherent limitation of studying a large number of participants using an extensive set of measurements, is that this provides information regarding the health status at that particular moment, and that the time between two measurement rounds is relatively long.

## Future research

HELIUS’s findings demonstrate the importance of studying diversity in the understanding of the etiology of disease as well as in improving health care and targeted prevention. Our results also underline the importance of addressing multiple risk factors at various spatial levels to reduce inequalities in health between ethnic groups. These factors range from living circumstances, such as number of people in the household during a pandemic, to social norms regarding obesity, to adverse childhood experiences, to environmental exposures such as air pollution, and to treatment adherence. Moreover, the results suggest that whereas improving the socioeconomic position of ethnic minority populations will lead to smaller inequalities in health (41), additional measures are needed, e.g. aimed at preventing discrimination.

In the years to come, we will continue with testing, refining and expanding our conceptual model. For example, the distinction between structural factors, group-level factors and individual-level factors can be emphasized more. The HELIUS follow-up data on incident diseases and their outcomes, with more certainty with regard to the temporal relationship and the avoidance of lead time bias, enriched by the analysis of repeated biological samples and the integration of data from health and other registries, could potentially aid in disentangling the influence of socioeconomic status and ethnicity in this context over time. Also, in order to obtain better insight in dynamics of diseases and their etiology, the adoption of different ways of data collection is explored, for example by applying ecological momentary assessment.

Future studies will focus on investigating the pathways for the observed differences in disease risk. This includes research on the underlying biological mechanisms such as changes in the microbiome, epigenome, and systemic inflammation associated with migration as well as creating reference standards and models that take ethnicity specific variations in biometric data into account. For example, HELIUS has shown that significant differences exist in normal values, for example for electrocardiogram criteria for diagnosing acute myocardial infarction (102). In addition, valid genome wide SNP arrays are currently not available for individuals of Surinamese and Moroccan descent populations. Further research also focuses on the synergy between the three research themes within HELIUS, which might bring new insights into improving health equity, and on finding prevention measures and clinical tools to mitigate differences in health and health care. Finally, we will further explore what factors explain inequalities *within* ethnic groups, thereby also focusing on examples of ‘positive deviance’: patterns in health that we did not expect, such as the lower risk of stroke among those of Moroccan descent (103), which might be a novel entry point for measures tackling health inequalities.

## Data availability

The HELIUS data are owned by the Amsterdam University Medical Centers, location AMC in Amsterdam, The Netherlands. Any researcher can request the data by submitting a proposal to the HELIUS Executive Board as outlined at http://www.heliusstudy.nl/en/researchers/collaboration, by. The HELIUS Executive Board will check proposals for compatibility with the general objectives, ethical approvals and informed consent forms of the HELIUS study. There are no other restrictions to obtaining the data and all data requests will be processed in the same manner. For linked data, additional conditions for reuse may apply. To allow sharing of microbiome data collected in HELIUS with (inter)national researchers, 16 s rRNA sequence analysis (accession codes EGAD00001004106, EGAD00001009732) and virome sequencing (accession code EGAD00001008765) have been stored at the European genome-phenome archive (EGA).

## Supplementary material

Supplementary Table 1. Comparing baseline demographic characteristics of groups with different follow-up status, by ethnic group.

Supplemental Table 2. Comparison of selected baseline health characteristics between participants who completed follow-up and those who did not. Excluding participants who deceased or moved abroad between baseline and follow-up.

Supplemental Table 3. Overview of COVID-related measurements in a subsample of the HELIUS study between 2020 and 2022.

Supplementary Figure 1. Demographic characteristics of HELIUS participants at HELIUS baseline and follow-up, by ethnic group.

## Acknowledgments

We are most grateful to the participants of the HELIUS study and the management team, research nurses, interviewers, research assistants and other staff who have taken part in gathering the data of this study. We also acknowledge the AMC biobank for their support in biobank management and high-quality storage of collected samples.

## Author contributions

All authors contributed to the study conception and design. HG and KS conceived the present manuscript. HG, AK, JZ, BB, MP and KS drafted the paper. HG and JZ performed the analyses. HG finalized writing the paper. AL, EMC, AV and AZ were involved in reviewing previous versions of the paper in detail. All authors read and approved the final manuscript.

## Ethics Approval

The HELIUS study is performed in line with the principles of the Declaration of Helsinki. The HELIUS study has been approved by the Ethical Review Board of the Academic Medical Center Amsterdam (Protocol ID NL32251.018.10; Approval numbers 10/100# 10.17.1729; 2010_100#B201997)

## Consent to participate

Written informed consent was obtained from all individual participants included in the study.

## Consent to publish

Not applicable.

## References

1. Flanagin A, Frey T, Christiansen SL, Committee AMoS. Updated Guidance on the Reporting of Race and Ethnicity in Medical and Science Journals. JAMA. 2021;326(7):621–7. doi:10.1001/jama.2021.13304

2. Bailey ZD, Feldman JM, Bassett MT. How Structural Racism Works — Racist Policies as a Root Cause of U.S. Racial Health Inequities. New England Journal of Medicine. 2020;384(8):768–73. doi:10.1056/NEJMms2025396

3. Stirbu I, Kunst AE, Bos V, Mackenbach JP. Differences in avoidable mortality between migrants and the native Dutch in The Netherlands. BMC Public Health. 2006;6(1):1–10.

4. Missinne S, Bracke P. Depressive symptoms among immigrants and ethnic minorities: a population based study in 23 European countries. Social Psychiatry and Psychiatric Epidemiology. 2012;47(1):97–109. doi:10.1007/s00127-010-0321-0

5. van Apeldoorn JAN, Agyemang C, Moll van Charante EP. Use of ethnic identifiers to narrow health inequality gaps. The Lancet Regional Health – Europe. 2022;18:100411. 10.1016/j.lanepe.2022.100411

6. Stronks K, Snijder MB, Peters RJ, Prins M, Schene AH, Zwinderman AH. Unravelling the impact of ethnicity on health in Europe: the HELIUS study. BMC Public Health. 2013;13(1):1–10. doi:10.1186/1471-2458-13-402

7. Stronks K, Kulu-Glasgow I, Agyemang C. The utility of ‘country of birth’ for the classification of ethnic groups in health research: the Dutch experience. Ethnicity & Health. 2009;14(3):255–69. doi:10.1080/13557850802509206

8. Snijder MB, Galenkamp H, Prins M, Derks EM, Peters RJG, Zwinderman AH, Stronks K. Cohort profile: the Healthy Life in an Urban Setting (HELIUS) study in Amsterdam, The Netherlands. BMJ Open. 2017;7(12). doi:10.1136/bmjopen-2017-017873

9. Ferwerda B, Abdellaoui A, Nieuwdorp M, Zwinderman K. A Genetic Map of the Modern Urban Society of Amsterdam. Front Genet. 2021;12:727269. doi:10.3389/fgene.2021.727269

10. Deschasaux M, Bouter KE, Prodan A, et al. Depicting the composition of gut microbiota in a population with varied ethnic origins but shared geography. Nat. Med. 2018;24(10):1526–31. doi:10.1038/s41591-018-0160-1

11. van der Vossen EW, Davids M, Bresser LR, et al. Gut microbiome transitions across generations in different ethnicities in an urban setting—the HELIUS study. Microbiome. 2023;11(1):99.

12. Borgdorff H, van der Veer C, van Houdt R, et al. The association between ethnicity and vaginal microbiota composition in Amsterdam, the Netherlands. PLoS One. 2017;12(7):e0181135. doi:10.1371/journal.pone.0181135

13. Hugenholtz F, van der Veer C, Terpstra ML, et al. Urine and vaginal microbiota compositions of postmenopausal and premenopausal women differ regardless of recurrent urinary tract infection and renal transplant status. Sci. Rep. 2022;12(1):2698 %@ 045-322.

14. Van de Wijgert JHHM, Verwijs MC, Gill AC, Borgdorff H, Van der Veer C, Mayaud P. Pathobionts in the vaginal microbiota: individual participant data meta-analysis of three sequencing studies. Frontiers in cellular and infection microbiology. 2020;10:129 %@ 2235-988.

15. Bos MM, Goulding NJ, Lee MA, et al. Investigating the relationships between unfavourable habitual sleep and metabolomic traits: evidence from multi-cohort multivariable regression and Mendelian randomization analyses. BMC Med. 2021;19(1):1–20.

16. Verhaar BJH, Mosterd CM, Collard D, et al. Sex differences in associations of plasma metabolites with blood pressure and heart rate variability: The HELIUS study. Atherosclerosis. 2023. doi:10.1016/j.atherosclerosis.2023.05.016

17. Gazzola K, Snijder MB, Hovingh GK, Stroes ESG, Peters RJG, van den Born BH. Ethnic differences in plasma lipid levels in a large multiethnic cohort: The HELIUS study. J. Clin. Lipidol. 2018;12(5):1217–24.e1. doi:10.1016/j.jacl.2018.06.015

18. van der Velden AIM, van den Berg BM, van den Born BJ, Galenkamp H, Ijpelaar DHT, Rabelink TJ. Ethnic differences in urinary monocyte chemoattractant protein-1 and heparanase-1 levels in individuals with type 2 diabetes: the HELIUS study. BMJ Open Diabetes Res Care. 2022;10(6). doi:10.1136/bmjdrc-2022-003003

19. Alberts CJ, Vos RA, Borgdorff H, et al. Vaginal high-risk human papillomavirus infection in a cross-sectional study among women of six different ethnicities in Amsterdam, the Netherlands: the HELIUS study. Sex. Transm. Infect. 2016;92(8):611–8. doi:10.1136/sextrans-2015-052397

20. Alberts CJ, Michel A, Bruisten S, Snijder MB, Prins M, Waterboer T, Schim van der Loeff MF. High-risk human papillomavirus seroprevalence in men and women of six different ethnicities in Amsterdam, the Netherlands: The HELIUS study. Papillomavirus Res. 2017;3:57–65. doi:10.1016/j.pvr.2017.01.003

21. Kovaleva A, Alberts CJ, Waterboer T, et al. A cross-sectional study on the concordance between vaginal HPV DNA detection and type-specific antibodies in a multi-ethnic cohort of women from Amsterdam, the Netherlands – the HELIUS study. BMC Infect. Dis. 2016;16(1):502. doi:10.1186/s12879-016-1832-4

22. Alberts CJ, Jeske R, de Martel C, et al. Helicobacter pylori seroprevalence in six different ethnic groups living in Amsterdam: The HELIUS study. Helicobacter. 2020;25(3):e12687. doi:10.1111/hel.12687

23. Alberts CJ, Schim van der Loeff MF, Sadik S, et al. Hepatitis E virus seroprevalence and determinants in various study populations in the Netherlands. PLoS One. 2018;13(12):e0208522. doi:10.1371/journal.pone.0208522

24. Zuure F, Bil J, Visser M, et al. Hepatitis B and C screening needs among different ethnic groups: A population-based study in Amsterdam, the Netherlands. JHEP Rep. 2019;1(2):71–80. doi:10.1016/j.jhepr.2019.04.003

25. Hulstein SH, Matser A, Alberts CJ, et al. Differences in Chlamydia trachomatis seroprevalence between ethnic groups cannot be fully explained by socioeconomic status, sexual healthcare seeking behavior or sexual risk behavior: a cross-sectional analysis in the HEalthy LIfe in an Urban Setting (HELIUS) study. BMC Infect. Dis. 2018;18(1):612. doi:10.1186/s12879-018-3533-7

26. Muilwijk M, Celis-Morales C, Nicolaou M, Snijder MB, Gill JMR, van Valkengoed IGM. Plasma Cholesteryl Ester Fatty Acids do not Mediate the Association of Ethnicity with Type 2 Diabetes: Results From the HELIUS Study. Mol. Nutr. Food Res. 2018;62(2). doi:10.1002/mnfr.201700528

27. Verhaar BJH, Collard D, Prodan A, et al. Associations between gut microbiota, faecal short-chain fatty acids, and blood pressure across ethnic groups: the HELIUS study. Eur. Heart J. 2020;41(44):4259–67. doi:10.1093/eurheartj/ehaa704

28. Muilwijk M, Callender N, Goorden S, Vaz FM, van Valkengoed IGM. Sex differences in the association of sphingolipids with age in Dutch and South-Asian Surinamese living in Amsterdam, the Netherlands. Biol. Sex Differ. 2021;12(1):13. doi:10.1186/s13293-020-00353-0

29. Muilwijk M, Goorden SMI, Celis-Morales C, et al. Contributions of amino acid, acylcarnitine and sphingolipid profiles to type 2 diabetes risk among South-Asian Surinamese and Dutch adults. BMJ Open Diabetes Res Care. 2020;8(1). doi:10.1136/bmjdrc-2019-001003

30. Fenneman AC, Boulund U, Collard D, et al. Compositional Disruption of the Gut Microbiota in a Multi-Ethnic Euthyroid Population with Thyroid Autoimmunity: The HELIUS Study. Available at SSRN 4309030.

31. Coyer L, Boyd A, Schinkel J, et al. Differences in SARS-CoV-2 infections during the first and second wave of SARS-CoV-2 between six ethnic groups in Amsterdam, the Netherlands: A population-based longitudinal serological study. Lancet Reg Health Eur. 2022;13:100284. doi:10.1016/j.lanepe.2021.100284

32. Chilunga FP, Coyer L, Collard D, et al. COVID-19 Impacts Across Multiple Life Domains of Vulnerable Socio-Demographic Groups Including Migrants: A Descriptive Cross-Sectional Study. Int J Public Health. 2022;67:1604665. doi:10.3389/ijph.2022.1604665

33. van Dijk AM, Vali Y, Mak AL, Galenkamp H, Nieuwdorp M, van den Born BJ, Holleboom AG. Noninvasive tests for nonalcoholic fatty liver disease in a multi-ethnic population: The HELIUS study. Hepatol Commun. 2023;7(1):e2109. doi:10.1002/hep4.2109

34. Vriend EMC, Bouwmeester TA, Va AA, et al. Ethnic Differences in Carotid Intima Media Thickness and Plaque Presence-The HELIUS study. Cerebrovascular Diseases (Basel, Switzerland) %@ 1015–9770. 2023.

35. Şekercan A, Snijder MB, Peters RJ, Stronks K. Associations between healthcare consumption in country of origin and in country of residence by people with Turkish and Moroccan backgrounds living in the Netherlands: the HELIUS study. Eur. J. Public Health. 2019;29(4):694–9. doi:10.1093/eurpub/ckz079

36. Schuts EC, van Dulm E, Boyd A, Snijder MB, Geerlings SE, Prins M, Prins JM. Knowledge and use of antibiotics in six ethnic groups: the HELIUS study. Antimicrob Resist Infect Control. 2019;8:200. doi:10.1186/s13756-019-0636-x

37. Bolijn R, Muilwijk M, Nicolaou M, et al. The contribution of smoking to differences in cardiovascular disease incidence between men and women across six ethnic groups in Amsterdam, the Netherlands: The HELIUS study. Prev Med Rep. 2023;31:102105. doi:10.1016/j.pmedr.2022.102105

38. van Tuijl LA, Voogd AC, de Graeff A, et al. Psychosocial factors and cancer incidence (PSY-CA): Protocol for individual participant data meta-analyses. Brain Behav. 2021;11(10):e2340. doi:10.1002/brb3.2340

39. van Tuijl LA, Basten M, Pan KY, et al. Depression, anxiety, and the risk of cancer: An individual participant data meta-analysis. Cancer. 2023. doi:10.1002/cncr.34853

40. Müller F, Veen LM, Galenkamp H, et al. Emotional distress in cancer survivors from various ethnic backgrounds: Analysis of the multi-ethnic HELIUS study. Psychooncology. 2023. doi:10.1002/pon.6192

41. Chilunga FP, Campman S, Galenkamp H, et al. Relative contributions of pre-pandemic factors and intra-pandemic activities to differential COVID-19 risk among migrant and non-migrant populations in the Netherlands: lessons for future pandemic preparedness. International Journal for Equity in Health. 2023;22(1):127. doi:10.1186/s12939-023-01936-0

42. Campman SL, Boyd A, Coyer L, et al. SARS-CoV-2 vaccination uptake in six ethnic groups living in Amsterdam, the Netherlands: A registry-based study within the HELIUS cohort. Preventive medicine. 2024;178:107822 %@ 0091-7435.

43. Generaal E, Hoogendijk EO, Stam M, et al. Neighbourhood characteristics and prevalence and severity of depression: pooled analysis of eight Dutch cohort studies. Br. J. Psychiatry. 2019;215(2):468–75. doi:10.1192/bjp.2019.100

44. Poelman MP, Nicolaou M, Dijkstra SC, et al. Does the neighbourhood food environment contribute to ethnic differences in diet quality? Results from the HELIUS study in Amsterdam, the Netherlands. Public Health Nutr. 2021;24(15):5101–12. doi:10.1017/s1368980021001919

45. Timmermans EJ, Veldhuizen EM, Snijder MB, Huisman M, Kunst AE. Neighbourhood safety and smoking in population subgroups: The HELIUS study. Prev. Med. 2018;112:111–8. doi:10.1016/j.ypmed.2018.04.012

46. Burger RJ, Gordijn SJ, Bolijn R, et al. Cardiovascular risk profile after a complicated pregnancy across ethnic groups: The HELIUS study. Eur J Prev Cardiol. 2022. doi:10.1093/eurjpc/zwac307

47. Snijder MB, Agyemang C, Peters RJ, Stronks K, Ujcic-Voortman JK, van Valkengoed IG. Case Finding and Medical Treatment of Type 2 Diabetes among Different Ethnic Minority Groups: The HELIUS Study. Journal of Diabetes Research. 2017;2017.

48. Agyemang C, Kieft S, Snijder MB, et al. Hypertension control in a large multi-ethnic cohort in Amsterdam, The Netherlands: the HELIUS study. Int. J. Cardiol. 2015;183:180–9. doi:10.1016/j.ijcard.2015.01.061

49. Agyemang C, Snijder MB, Adjei DN, et al. Ethnic Disparities in CKD in the Netherlands: The Healthy Life in an Urban Setting (HELIUS) Study. Am. J. Kidney Dis. 2016;67(3):391–9. doi:10.1053/j.ajkd.2015.07.023

50. Armengol GD, Hayfron-Benjamin CF, van den Born BH, Galenkamp H, Agyemang C. Microvascular and macrovascular complications in type 2 diabetes in a multi-ethnic population based in Amsterdam. The HELIUS study. Prim. Care Diabetes. 2021;15(3):528–34. doi:10.1016/j.pcd.2021.02.008

51. Perini W, Kunst AE, Snijder MB, Peters RJG, van Valkengoed IGM. Ethnic differences in metabolic cardiovascular risk among normal weight individuals: Implications for cardiovascular risk screening. The HELIUS study. Nutr. Metab. Cardiovasc. Dis. 2019;29(1):15–22. doi:10.1016/j.numecd.2018.09.004

52. Zethof M, Mosterd CM, Collard D, et al. Differences in Body Composition Convey a Similar Risk of Type 2 Diabetes Among Different Ethnic Groups With Disparate Cardiometabolic Risk-The HELIUS Study. Diabetes Care. 2021;44(7):1692–8. doi:10.2337/dc21-0230

53. NHG-werkgroep:: Barents ESE BH, Bouma M, Dankers M, De Rooij A, Hart HE, Houweling ST, IJzerman RG, Janssen PGH, Kerssen A, Oud M, Palmen J, Van den Brink-Muinen A, Van den Donk M, Verburg-Oorthuizen AFE, Wiersma Tj NHG-Standaard Diabetes mellitus type 2 (M01). Versie 5.6. 2023.

54. Agyemang C, Kieft S, Snijder MB, et al. Hypertension control in a large multi-ethnic cohort in Amsterdam, The Netherlands: The HELIUS study. Int. J. Cardiol. 2015;183:180–9.

55. van Laer SD, Snijder MB, Agyemang C, Peters RJ, van den Born BH. Ethnic differences in hypertension prevalence and contributing determinants – the HELIUS study. Eur J Prev Cardiol. 2018;25(18):1914–22. doi:10.1177/2047487318803241

56. Blok S, Haggenburg S, Collard D, et al. The association between socioeconomic status and prevalence, awareness, treatment and control of hypertension in different ethnic groups: the Healthy Life in an Urban Setting study. J. Hypertens. 2022;40(5):897–907. doi:10.1097/hjh.0000000000003092

57. Kist JM, Vos RC, Mairuhu ATA, et al. SCORE2 cardiovascular risk prediction models in an ethnic and socioeconomic diverse population in the Netherlands: an external validation study. (2589-5370 (Electronic)).

58. Meinema JG, van Dijk N, Beune EJ, Jaarsma DA, van Weert HC, Haafkens JA. Determinants of adherence to treatment in hypertensive patients of African descent and the role of culturally appropriate education. (1932-6203 (Electronic)).

59. Perini W, Agyemang C, Snijder MB, Peters RJG, Kunst AE. Ethnic disparities in treatment rates for hypertension and dyslipidemia: an analysis by different treatment indications: the Healthy Life in an Urban Setting study. J. Hypertens. 2018;36(7):1540–7. doi:10.1097/hjh.0000000000001716

60. Vriend EMC, Wever BE, Bouwmeester TA, et al. Ethnic Differences in Blood Pressure Levels over Time – The HELIUS Study. Eur J Prev Cardiol. 2023. doi:10.1093/eurjpc/zwad089

61. Perini W, van Valkengoed IGM, Snijder MB, Peters RJG, Kunst AE. The contribution of obesity to the population burden of high metabolic cardiovascular risk among different ethnic groups. The HELIUS study. Eur. J. Public Health. 2020;30(2):322–7. doi:10.1093/eurpub/ckz190

62. Ikram UZ, Snijder MB, Fassaert TJL, Schene AH, Kunst AE, Stronks K. The contribution of perceived ethnic discrimination to the prevalence of depression. The European Journal of Public Health. 2015;25(2):243–8. doi:10.1093/eurpub/cku180

63. Stronks K, Şekercan A, Snijder M, Lok A, Verhoeff AP, Kunst AE, Galenkamp H. Higher prevalence of depressed mood in immigrants’ offspring reflects their social conditions in the host country: The HELIUS study. PLoS One. 2020;15(6):e0234006. doi:10.1371/journal.pone.0234006

64. van Amsterdam J, Vorspan F, Snijder MB, et al. Use of the Fagerström test to assess differences in the degree of nicotine dependence in smokers from five ethnic groups: The HELIUS study. Drug Alcohol Depend. 2019;194:197–204. doi:10.1016/j.drugalcdep.2018.10.011

65. Miller AP, Merkle EC, Galenkamp H, Stronks K, Derks EM, Gizer IR. Differential item functioning analysis of the CUDIT and relations with alcohol and tobacco use among men across five ethnic groups: The HELIUS study. Psychol. Addict. Behav. 2019;33(8):697–709. doi:10.1037/adb0000521

66. Ikram UZ, Snijder MB, Derks EM, Peters RJG, Kunst AE, Stronks K. Parental Smoking and Adult Offspring’s Smoking Behaviors in Ethnic Minority Groups: An Intergenerational Analysis in the HELIUS Study. Nicotine Tob Res. 2018;20(6):766–74. doi:10.1093/ntr/ntx137

67. van Leijden MJ, Penninx B, Agyemang C, Olff M, Adriaanse MC, Snijder MB. The association of depression and posttraumatic stress disorder with the metabolic syndrome in a multi-ethnic cohort: the HELIUS study. Soc Psychiatry Psychiatr Epidemiol. 2018;53(9):921–30. doi:10.1007/s00127-018-1533-y

68. Willemen FEM, Heuschen C, Zantvoord JB, et al. Perceived ethnic discrimination, suicidal ideation and mastery in a multi-ethnic cohort: the HELIUS study. BJPsych Open. 2023;9(1):e21. doi:10.1192/bjo.2022.640

69. van Amsterdam JGC, Benschop A, van Binnendijk S, et al. A Comparison of Excessive Drinking, Binge Drinking and Alcohol Dependence in Ethnic Minority Groups in the Netherlands: The HELIUS Study. Eur. Addict. Res. 2020;26(2):66–76. doi:10.1159/000504881

70. Sunley AK, Lok A, White MJ, Snijder MB, van Zuiden M, Zantvoord JB, Derks EM. Ethnic and sex differences in the association of child maltreatment and depressed mood. The HELIUS study. Child Abuse Negl. 2020;99:104239. doi:10.1016/j.chiabu.2019.104239

71. Willemen FEM, van Zuiden M, Zantvoord JB, et al. Associations Between Child Maltreatment, Inflammation, and Comorbid Metabolic Syndrome to Depressed Mood in a Multiethnic Urban Population: The HELIUS Study. Front. Psychol. 2022;13:787029. doi:10.3389/fpsyg.2022.787029

72. Slotman A, Snijder MB, Ikram UZ, Schene AH, Stevens GW. The role of mastery in the relationship between perceived ethnic discrimination and depression: The HELIUS study. Cultur. Divers. Ethnic Minor. Psychol. 2017;23(2):200–8. doi:10.1037/cdp0000113

73. Hagen JM, Sutterland AL, Collard D, et al. Ethnic discrimination and depressed mood: The role of autonomic regulation. J. Psychiatr. Res. 2021;144:110–7. doi:10.1016/j.jpsychires.2021.09.048

74. Vermeulen E, Stronks K, Snijder MB, et al. A combined high-sugar and high-saturated-fat dietary pattern is associated with more depressive symptoms in a multi-ethnic population: the HELIUS (Healthy Life in an Urban Setting) study. Public Health Nutr. 2017;20(13):2374–82. doi:10.1017/s1368980017001550

75. Vermeulen E, Stronks K, Visser M, et al. Dietary pattern derived by reduced rank regression and depressive symptoms in a multi-ethnic population: the HELIUS study. Eur J Clin Nutr. 2017;71(8):987–94. doi:10.1038/ejcn.2017.61

76. Nicolaou M, Colpo M, Vermeulen E, et al. Association of a priori dietary patterns with depressive symptoms: a harmonised meta-analysis of observational studies. Psychol. Med. 2020;50(11):1872–83. doi:10.1017/s0033291719001958

77. Anujuo K, Stronks K, Snijder MB, Lok A, Jean-Louis G, Agyemang C. Association between Depressed Mood and Sleep Duration among Various Ethnic Groups-The Helius Study. Int. J. Environ. Res. Public Health. 2021;18(13). doi:10.3390/ijerph18137134

78. Brathwaite R, Smeeth L, Addo J, et al. Ethnic differences in current smoking and former smoking in the Netherlands and the contribution of socioeconomic factors: a cross-sectional analysis of the HELIUS study. BMJ Open. 2017;7(7):e016041. doi:10.1136/bmjopen-2017-016041

79. Nieuwenhuijsen K, Schene AH, Stronks K, Snijder MB, Frings-Dresen MH, Sluiter JK. Do unfavourable working conditions explain mental health inequalities between ethnic groups? Cross-sectional data of the HELIUS study. BMC Public Health. 2015;15:805. doi:10.1186/s12889-015-2107-5

80. Leijssen JB, Snijder MB, Timmermans EJ, Generaal E, Stronks K, Kunst AE. The association between road traffic noise and depressed mood among different ethnic and socioeconomic groups. The HELIUS study. Int. J. Hyg. Environ. Health. 2019;222(2):221–9. doi:10.1016/j.ijheh.2018.10.002

81. Gibson-Smith D, Bot M, Snijder M, et al. The relation between obesity and depressed mood in a multi-ethnic population. The HELIUS study. Soc Psychiatry Psychiatr Epidemiol. 2018;53(6):629–38. doi:10.1007/s00127-018-1512-3

82. Fernald F, Snijder M, van den Born BJ, Lok A, Peters R, Agyemang C. Depression and hypertension awareness, treatment, and control in a multiethnic population in the Netherlands: HELIUS study. Intern. Emerg. Med. 2021;16(7):1895–903. doi:10.1007/s11739-021-02717-9

83. Bosch JA, Nieuwdorp M, Zwinderman AH, et al. The gut microbiota and depressive symptoms across ethnic groups. Nat Commun. 2022;13(1):7129. doi:10.1038/s41467-022-34504-1

84. Radjabzadeh D, Bosch JA, Uitterlinden AG, et al. Gut microbiome-wide association study of depressive symptoms. Nat Commun. 2022;13(1):7128. doi:10.1038/s41467-022-34502-3

85. Netherlands HCot. Screening Risk Groups for Hepatitis B and C. The Hague: Health Council of the Netherlands; Publication No. 2016/16. 2016.

86. Minister of Health WaSitN. Geneesmiddelenbeleid. In: Health WaS, editor. Den Haag, The Netherlands: Tweede Kamer der Staten Generaal; 2017.

87. Edouard Mathieu HR, Lucas Rodés-Guirao, Cameron Appel, Charlie Giattino, Joe Hasell, Bobbie Macdonald, Saloni Dattani, Diana Beltekian, Esteban Ortiz-Ospina and Max Roser. Coronavirus Pandemic (COVID-19). OurWorldInData.org. 2020. https://ourworldindata.org/coronavirus. 2024.

88. Collard D, Stronks K, Harris V, et al. Ethnic Differences in Coronavirus Disease 2019 Hospitalization and Hospital Outcomes in a Multiethnic Population in the Netherlands. Open Forum Infect Dis. 2022;9(6):ofac257. doi:10.1093/ofid/ofac257

89. Campman SL, van Rossem G, Boyd A, et al. Intent to vaccinate against SARS-CoV-2 and its determinants across six ethnic groups living in Amsterdam, the Netherlands: A cross-sectional analysis of the HELIUS study. Vaccine. 2023;41(12):2035–45. doi:10.1016/j.vaccine.2023.02.030

90. Read JnG, Emerson MO, Tarlov A. Implications of black immigrant health for US racial disparities in health. Journal of Immigrant Health. 2005;7:205–12.

91. Perini W, Snijder MB, Peters RJG, Stronks K, Kunst AE. Increased cardiovascular disease risk in international migrants is independent of residence duration or cultural orientation: the HELIUS study. J Epidemiol Community Health. 2018;72(9):825–31. doi:10.1136/jech-2018-210595

92. Amoah AS, Prins M, Bel EHD, et al. Migration and allergic diseases: Findings from a population-based study in adults in Amsterdam, the Netherlands. Allergy. 2022;77(12):3667–70. doi:10.1111/all.15427

93. Balvers M, Deschasaux M, van den Born BJ, Zwinderman K, Nieuwdorp M, Levin E. Analyzing Type 2 Diabetes Associations with the Gut Microbiome in Individuals from Two Ethnic Backgrounds Living in the Same Geographic Area. Nutrients. 2021;13(9). doi:10.3390/nu13093289

94. Crielaard L, Dutta P, Quax R, Nicolaou M, Merabet N, Stronks K, Sloot PMA. Social norms and obesity prevalence: from cohort to system dynamics models. Obes. Rev. 2020;21(9):e13044 %@ 1467-7881.

95. Habibe JJ, Boulund U, Clemente-Olivo MP, et al. FHL2 Genetic Polymorphisms and Pro-Diabetogenic Lipid Profile in the Multiethnic HELIUS Cohort. Int. J. Mol. Sci. 2023;24(5). doi:10.3390/ijms24054332

96. Boulund U, Bastos DM, Ferwerda B, et al. Gut microbiome associations with host genotype vary across ethnicities and potentially influence cardiometabolic traits. Cell Host Microbe. 2022;30(10):1464–80.e6. doi:10.1016/j.chom.2022.08.013

97. Sturkenboom SM, Dekker LH, Lamkaddem M, Schaap LA, de Vries JH, Stronks K, Nicolaou M. Acculturation and dietary patterns among residents of Surinamese origin in the Netherlands: the HELIUS dietary pattern study. Public Health Nutr. 2016;19(4):682–92. doi:10.1017/s1368980015001391

98. Hoenink JC, Galenkamp H, Beune EJ, et al. Body size ideals and body satisfaction among Dutch-origin and African-origin residents of Amsterdam: The HELIUS study. PLoS One. 2021;16(5):e0252054. doi:10.1371/journal.pone.0252054

99. Galenkamp H, Stronks K, Mokkink LB, Derks EM. Measurement invariance of the SF-12 among different demographic groups: The HELIUS study. PLoS One. 2018;13(9):e0203483. doi:10.1371/journal.pone.0203483

100. Galenkamp H, Stronks K, Snijder MB, Derks EM. Measurement invariance testing of the PHQ-9 in a multi-ethnic population in Europe: the HELIUS study. BMC Psychiatry. 2017;17(1):349. doi:10.1186/s12888-017-1506-9

101. Souama C, Lamers F, Milaneschi Y, et al. Depression, cardiometabolic disease, and their co-occurrence after childhood maltreatment: an individual participant data meta-analysis including over 200,000 participants. BMC Med. 2023;21(1):93. doi:10.1186/s12916-023-02769-y

102. Ter Haar CC, Kors JA, Peters RJG, et al. Prevalence of ECGs Exceeding Thresholds for ST-Segment-Elevation Myocardial Infarction in Apparently Healthy Individuals: The Role of Ethnicity. J Am Heart Assoc. 2020;9(13):e015477. doi:10.1161/jaha.119.015477

103. Agyemang C, van Oeffelen AA, Norredam M, et al. Socioeconomic inequalities in stroke incidence among migrant groups: analysis of nationwide data. Stroke. 2014;45(8):2397–403. doi:10.1161/strokeaha.114.005505

