## Supplementary material for "The Healthy Life in an Urban Setting (HELIUS) study in Amsterdam, The Netherlands: cohort update 2024 and key findings"

Supplementary Table 1. Comparing baseline demographic characteristics of groups with different follow-up status, by ethnic group.

|  | **Total*** | **Dutch** | **South-Asian Surinamese** | **African Surinamese** | **Ghanaian** | **Turkish** | **Moroccan** |
| --- | --- | --- | --- | --- | --- | --- | --- |
| Sex (% women) |  |  |  |  |  |  |  |
| Baseline sample | 57.4 | 54.1 | 53.7 | 59.5 | 61.1 | 54.3 | 61.9 |
| Follow-up sample | 56.7 | 52.3 | 56.5 | 63.4 | 60.7 | 52.3 | 56.3 |
| Deceased | 36.7 | 36.0 | 32.9 | 37.0 | 40.5 | 31.0 | 54.2 |
| Moved abroad | 51.1 | 50.0 | 53.8 | 44.1 | 55.6 | 47.9 | 58.5 |
| Refused | 59.5 | 58.8 | 53.9 | 61.9 | 64.9 | 55.2 | 64.6 |
| No contact | 58.2 | 58.1 | 46.3 | 53.3 | 60.1 | 56.8 | 66.2 |
| Age (years (sd)) |  |  |  |  |  |  |  |
| Baseline sample | 43.8 (13.4) | 46.1 (14.1) | 45.1 (13.5) | 47.6 (12.8) | 44.0 (11.7) | 39.9 (12.5) | 39.7 (13.1) |
| Follow-up sample | 46.3 (12.4) | 47.7 (13.1) | 46.6 (12.4) | 49.5 (11.5) | 45.6 (10.2) | 41.4 (11.4) | 42.4 (12.1) |
| Deceased | 56.5 (10.4) | 58.9 (9.5) | 57.3 (9.6) | 58.0 (9.2) | 54.4 (10.2) | 51.8 (12.8) | 51.5 (13.0) |
| Moved abroad | 39.8 (13.7) | 37.7 (13.0) | 42.1 (13.1) | 43.6 (14.4) | 37.7 (13.0) | 40.7 (13.4) | 35.8 (14.1) |
| Refused | 43.0 (13.6) | 44.7 (15.1) | 44.7 (14.4) | 47.4 (13.3) | 44.5 (11.6) | 39.9 (12.4) | 39.8 (13.0) |
| No contact | 39.4 (13.5) | 34.7 (12.6) | 39.1 (13.7) | 43.4 (13.3) | 42.5 (12.5) | 37.7 (13.1) | 36.5 (13.5) |
| Migration generation (% 1st generation) |  |  |  |  |  |  |  |
| Baseline sample | 76.6 | - | 75.5 | 82.8 | 94.4 | 68.7 | 66.6 |
| Follow-up sample | 81.8 | - | 80.7 | 85.4 | 96.4 | 73.5 | 74.6 |
| Deceased | 95.3 | - | 94.7 | 97.0 | 100 | 88.1 | 91.7 |
| Moved abroad | 78.2 | - | 71.2 | 83.5 | 90.7 | 70.8 | 60.4 |
| Refused | 74.3 | - | 71.9 | 79.5 | 94.2 | 69.1 | 66.3 |
| No contact | 70.0 | - | 62.1 | 78.9 | 92.9 | 62.0 | 57.3 |
| Educational level (% elementary or lower secondary/vocational education)** |  |  |  |  |  |  |  |
| Baseline sample | 44.3 | 17.6 | 47.6 | 41.9 | 68.0 | 56.3 | 48.6 |
| Follow-up sample | 38.0 | 14.9 | 45.7 | 37.8 | 68.7 | 48.9 | 48.6 |
| Deceased | 64.6 | 35.2 | 69.3 | 62.2 | 84.6 | 82.9 | 83.3 |
| Moved abroad | 42.4 | 8.5 | 29.4 | 44.1 | 54.5 | 57.8 | 43.1 |
| Refused | 47.7 | 24.4 | 50.1 | 43.0 | 67.0 | 57.1 | 48.8 |
| No contact | 52.0 | 15.6 | 47.9 | 47.1 | 69.5 | 61.5 | 47.9 |

*Including 853 participants from other ethnic backgrounds; **Excluding n=333 with unknown educational level

Supplemental Table 2. Comparison of selected baseline health characteristics between participants who completed follow-up and those who did not. Excluding participants who deceased or moved abroad between baseline and follow-up.

|  |  | Total* | Dutch | South-Asian Surinamese | African Surinamese | Ghanaian | Turkish | Moroccan |
| --- | --- | --- | --- | --- | --- | --- | --- | --- |
| Diabetes (%) | **H2** | 9.3 | 3.2 | 17.9 | 10.1 | 9.8 | 8.0 | 10.8 |
|  | **No H2** | 11.9 | 4.3 | 20.5 | 13.6 | 12.3 | 10.5 | 11.7 |
|  | **p-value**** | <.001 | 0.099 | 0.085 | <.001 | 0.063 | 0.020 | 0.388 |
| Hypertension (%) | **H2** | 37.3 | 29.6 | 41.1 | 50.7 | 54.4 | 27.5 | 26.2 |
|  | **No H2** | 36.7 | 28.8 | 43.5 | 49.8 | 56.3 | 29.3 | 22.7 |
|  | **p-value** | 0.380 | 0.572 | 0.209 | 0.587 | 0.397 | 0.298 | 0.012 |
| Self-reported CVD (%) | **H2** | 4.5 | 3.1 | 7.5 | 4.9 | 4.0 | 5.5 | 3.3 |
|  | **No H2** | 5.9 | 4.7 | 10.9 | 5.7 | 5.0 | 6.9 | 3.3 |
|  | **p-value** | <.001 | 0.010 | 0.001 | 0.271 | 0.298 | 0.125 | 1.000 |
| Current smoker (%) | **H2** | 21.1 | 22.0 | 24.0 | 27.9 | 4.9 | 29.6 | 10.3 |
|  | **No H2** | 27.8 | 30.5 | 37.6 | 37.4 | 3.8 | 37.7 | 15.9 |
|  | **p-value** | <.001 | <.001 | <.001 | <.001 | 0.238 | <.001 | <.001 |
| Alcohol use (intermediate/high ***) (%) | H2 | **27.1** | **64.8** | **15.7** | **18.3** | **11.5** | **8.2** | **3.3** |
|  | No H2 | 16.5 | 59.6 | 17.8 | 20.2 | 10.0 | 5.8 | 3.1 |
|  | p-value | <.001 | <.001 | 0.106 | 0.117 | 0.260 | 0.007 | 0.720 |
| Meeting Dutch physical activity guideline (%) | **H2** | 59.6 | 75.5 | 53.1 | 61.3 | 54.7 | 43.3 | 47.8 |
|  | **No H2** | 46.0 | 72.1 | 42.9 | 52.8 | 48.0 | 34.5 | 38.5 |
|  | **p-value** | <.001 | 0.016 | <.001 | <.001 | 0.002 | <.001 | <.001 |
| Depressive symptoms (PHQ-9 sum score >=10) (%) | **H2** | 12.1 | 6.0 | 17.3 | 9.2 | 8.8 | 20.5 | 18.7 |
|  | **No H2** | 17.0 | 9.3 | 20.1 | 12.0 | 9.1 | 23.4 | 21.5 |
|  | **p-value** | <.001 | <.001 | 0.050 | 0.004 | 0.820 | 0.054 | 0.033 |

**including other ethnic backgrounds; ** chi-square test (2 sided exact test);*** Intermediate/high (men >4 glasses/week, women >2 glasses/week)
H2: participation in HELIUS-2; No H2: no participation in HELIUS-2 (because of refusal or no contact)*

Supplemental Table 3. Overview of COVID-related measurements in a subsample of the HELIUS study between 2020 and 2022.

|  | **HELIUS-COVID-1**  **June 2020- Oct 2020** | **HELIUS-COVID-2**  **Nov 2020- June 2021** | **HELIUS-COVID-3**  **May 2022- Nov 2022** |
| --- | --- | --- | --- |
|  | **N=2496** | **N=2088** | **N=1505** |
| SARS-CoV-2 antibody test result | **X** | **X** | **X** |
| *Self-report of:* |  |  |  |
| SARS-CoV-2 testing behaviors (tested since start 2020 or previous round, test type, how often, test location) | **X** | **X** | **X** |
| Testing intention and motivation |  |  | **X** |
| Possibility infection participant (confirmed y/n, date) | **X** | **X** | **X** |
| COVID-19-related symptoms | **X** | **X** | **X** |
| Duration COVID-19-related symptoms |  |  | **X** |
| Hospitalized for COVID-19 | **X** | **X** | **X** |
| Trauma after hospitalization for COVID-19 | **X** | **X** |  |
| During/after infection: Working environment, sick leave, duration absence |  |  | **X** |
| General health, symptoms, fatigue, ability to work |  |  | **X** |
| Household members (age range and relation) | **X** |  |  |
| Possibility infection household member/steady partner/close contact | **X** | **X** | **X** |
| Household member/close contact hospitalized for COVID-19 | **X** | **X** | **X** |
| Work environment | **X** |  | **X** |
| Traveling abroad in 2020 or since previous round | **X** | **X** |  |
| COVID-19 behaviors in the past week (i.e. number of times leaving the house, type of locations visited, keeping distance, number and frequency of visitors, frequency of using public transportation) | **X** | **X** |  |
| Sources for information regarding COVID-19 | **X** |  |  |
| Vaccination-related variables (i.e. target audience for flu vaccination, vaccinated y/n and when for flu, COVID or TB, intention to get COVID vaccination) |  | **X** |  |
| Vaccinated for COVID y/n, number of doses, motivation, date last dose, intention to get new dose |  |  | **X** |
| Vitamin D intake |  | **X** |  |
| Perceived discrimination with regard to COVID |  | **X** |  |
| Perception corona virus |  | **X** |  |
| Memory, concentration, perception COVID measures |  |  | **X** |
| Mental health (PHQ-9) | **X** | **X** | **X** |

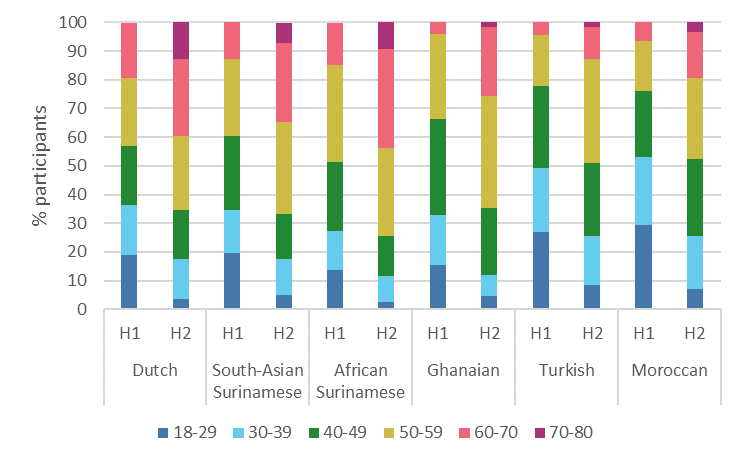

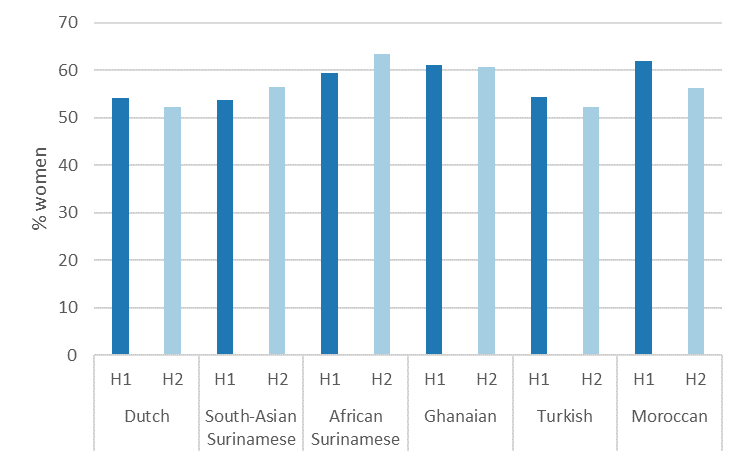

1B

1 A

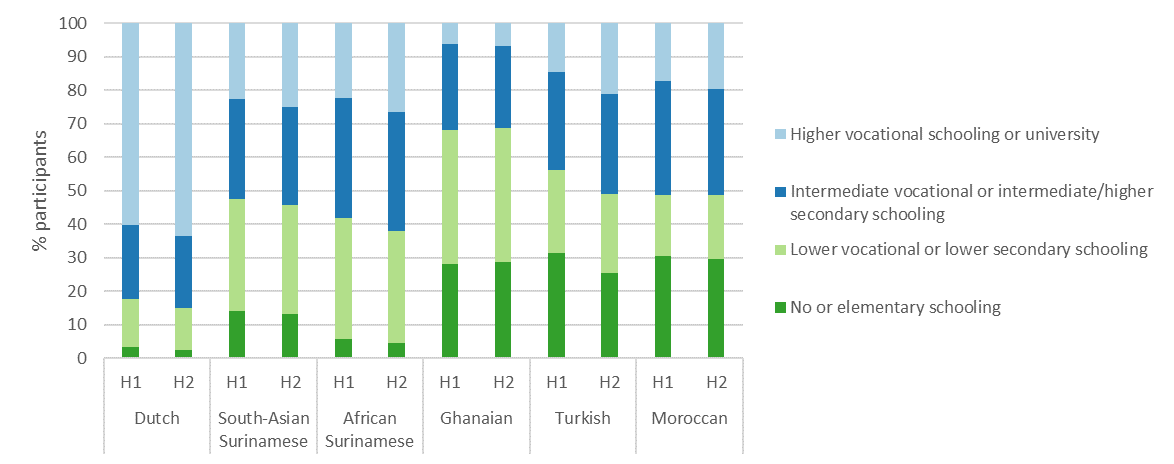

1 C

Supplementary Figure 1. Demographic characteristics of HELIUS participants at HELIUS baseline (H1) and follow-up (H2), by ethnic group.
1 A: Percentage of women; 1 B: Percentage of participants in each age group; 1 C: Percentage of participants within each educational level.
